# Analyzing Covid-19 Data using SIRD Models

**DOI:** 10.1101/2020.05.28.20115527

**Authors:** Abhijit Chakraborty, Jiaying Chen, Amélie Desvars-Larrive, Peter Klimek, Erwin Flores Tames, David Garcia, Leonhard Horstmeyer, Michaela Kaleta, Jana Lasser, Jenny Reddish, Beate Pinior, Johannes Wachs, Peter Turchin

## Abstract

The goal of this analysis is to estimate the effects of the diverse government intervention measures implemented to mitigate the spread of the Covid-19 epidemic. We use a process model based on a compartmental epidemiological framework Susceptible-Infected-Recovered-Dead (SIRD). Analysis of case data with such a mechanism-based model has advantages over purely phenomenological approaches because the parameters of the SIRD model can be calibrated using prior knowledge. This approach can be used to investigate how governmental interventions have affected the Covid-19-related transmission and mortality rate during the epidemic.

## Introduction

An outbreak of “pneumonia of unknown etiology” detected in Wuhan, Hubei Province, China in early December 2019 has induced a large epidemic in China (Li et al. 2020) and has become a worldwide threat, as it was declared a global pandemic by the World Health Organization (WHO) (WHO Regional Office for Europe 2020). The causative, newly emerging, agent was identified as a new betacoronavirus and named Severe Acute Respiratory Syndrome virus coronavirus 2 (SARS-Cov2), as it is genetically related to the SARS-CoV1 that emerged in 2003. The SARS-Cov2 associated disease has been named “COVID-19” by the WHO on 11 February 2020 (WHO 2020l).

Similarly, to the SARS-CoV1 and Middle East Respiratory Syndrome (MERS) coronavirus, the SARS-Cov2 is transmitted from person to person principally by respiratory droplets and induces, in most cases, fever and respiratory symptoms, such as cough, and shortness of breath. The incubation time (latent period) is estimated to be between 2 and 14 days. To date few cases have been documented in children (Z. Wu and McGoogan 2020; Cai et al. 2020).

China implemented very stringent intervention measures in response to the outbreak. On January 23, 2020, a cordon sanitaire was set around Wuhan (known as the “Wuhan lockdown”). In the following week, the traffic was suspended, a home quarantine was enforced, all gatherings were prohibited, and wearing a face mask in public place became mandatory (Berlinger, George, and Kottasová 2020; Cyranoski and Silver 2020). Despite these draconian intervention measures, the epidemic continued to spread, with the number of new cases per day reaching a peak in early February and the epidemic was controlled only in March (for a detailed discussion of Covid-19 in Wuhan, see section *Hubei, China* below). Later, worldwide, countries have implemented unprecedented mitigating interventions, before the introduction of the disease on their respective territories and in response to the increasing number of cases. However, how those interventions affected the transmission potential of the Covid-19 is still largely unknown.

Predictive mathematical models for epidemics are essential to understand the course of the epidemic and to inform preparedness and control strategy plans, as the recent Ebola epidemic made clear (Merler et al. 2015). A paradigmatic mathematical approach to describe and forecast the course of an epidemic is so-called compartmental models (Brauer 2008). In the simplest case of such models, the population is segmented into three different subpopulations or compartments: **S**usceptible, **I**nfected and **R**ecovered (SIR) individuals (Kermack, McKendrick, and Walker 1927). There is a constant rate at which susceptibles get infected after a contact; infected people recover with a constant rate. In these models, epidemic outbreaks only end once the susceptible population has been depleted to a point where one infected on average transmits the disease to less than one other person, the threshold of “herd immunity.”

There is a bestiary of model variations in which additional compartments are added to the SIR baseline, such as an exposed state of individuals that have been infected but are not yet infectious themselves, giving a so-called SEIR model (Liu, Hethcote, and Levin 1987). If individuals become susceptible again after recovering, one has a SEIRS model, and so forth. A wide range of epidemic growth patterns can be explained by such models by including more spatial details, more complex population structures, and how different subpopulations mix (Chowell et al. 2016). For instance, in many countries the “flattening of the curve” of the COVID-19 epidemic can be described by extensions of SIR models that model the isolation and quarantine of infected individuals, as well as lockdown and social distancing measures imposed on the general population (Maier and Brockmann 2020). Recent years also brought a surge in epidemic models in which susceptible and infected people do not meet randomly (as in SIR-type models) but interact on social networks (Keeling and Eames 2005), giving rise to sophisticated agent-based modeling approaches for epidemic forecasts (Ajelli et al. 2010; Balcan et al. 2010).

Here, we use an SIRD model in which individuals either recover or die after being infected. We differ from the standard textbook formulation of compartmental models by assuming that the rates at which individuals transition between compartments changes dynamically. These time-dependent rates are often account for seasonality effects (Stone, Olinky, and Huppert 2007). In this study, we assume that the time-dependence is a direct consequence of non-pharmaceutical intervention measures. Therefore, the effectiveness of these interventions can be estimated by measuring the strength of the concurrent reduction in disease transmission rates.

In this research paper, we propose a model that examine the course of the Covid-19 epidemic and applied it to 29 country case studies. The model considers four stages of infection: susceptible (S), infected (I), recovered (R), and dead (D).

## Materials and Methods

### Modeling Framework

The starting point is a discrete-time version of a standard compartmental epidemiological SIRD (susceptible–infected–recovered–dead) model.

#### Variables

*t* time in days
*S*(*t*) the number of susceptible at time *t*
*I*(*t*) the number of infected at time *t*
*R*(*t*) the number of recovered at time *t*
*D*(*t*) the number of deaths at time *t*
*N* = *S* + *I* + *R* + *D*, the total population (approximately a constant)

#### *Equations* (*suppressing time-dependence on the right-hand side, thus, S*(*t*) *→ S*)

*S*(*t+*1) = *S* − *βSI/N*
*I*(*t+*1) = *I* + *βSI/N* − *γI* − *δI*
*R*(*t+*1) = *R* + *γI*
*D*(*t*+1) = *D* + *δI*

#### Parameters

β transmission coefficient
γ recovery rate
δ death rate

The exponential growth rate during the initial phase of the epidemic, when *S ≈ N*, is *r*_0_ = β − (γ + δ). Note that this is the (per capita) growth rate per day; thus, its unit is day^−1^. It is different from the basic reproductive number, *R*_0_ = β/(γ + δ) which is dimensionless. Epidemic grows when *r*_0_ > 0 (*R*_0_ > 1) and dies out otherwise.

#### Estimating Intervention Impact on SIRD Parameters

Let ∆*S = S*(*t*+1) − *S*(*t*) be the daily change in *S*. ∆*I*, ∆*R*, ∆*D* are defined analogously. Rewriting the discrete SIRD models in the differenced form we obtain

∆*S* = − β*SI/N*

∆*I* = β*SI/N* − γ*I* − δ*I*

∆*R* = γ*I*

∆*D* = *δI*

Solving these equations for parameters we have

β = ∆*C/I*
γ = ∆*R/I*
δ = ∆*D/I*

where ∆*C* is change per day in the total number of cases, *C* = *I* + *R* + *D*. We also assumed that *S ≈ N* (because the number of total cases so far has been less than 1 percent of the total population).

Thus, if we have data on daily change in *I*, *R*, and *D* we can obtain sequential estimates of the SIRD parameters on a daily basis. Naturally, these estimates will be very noisy due to both process and observation noise. While the number of infected are relatively small at the beginning of the pandemic, the process noise is expected to be particularly high. Measurement noise can arise as a result of a number of factors. First, there can be delays in reporting cases. A constant delay causes no problem for this analysis, because it merely shifts all trajectories by a constant time lag. However, the delay in reporting may also change. For example, if no data are reported on a weekend, we will observe zeros infected over the weekend and a cumulative number (over three days) on Monday. Second, we expect that the detection rate of new cases will be low in the beginning of the epidemic and then gradually increase as the awareness of the epidemic spreads. Because many individuals infected with SARS-CoV-2 are asymptomatic, the prevalence of testing asymptomatic cases will affect how many cases are reported. This affects both new cases and deaths, because initially many deaths due to Covid-19 may be misclassified as pneumonia caused by other diseases. Third, government agencies responsible for collecting data may change their definitions of what constitutes “infected” and “recovered”.

All these sources of measurement error will tend to obscure the changes in epidemiological rates resulting from interventions. Smoothing the daily estimates helps to reveal patterns in the data. This is an effective procedure for reducing the error arising from variable reporting delays. Ultimately the results reported below are informative only as long as we can filter out the effect of various errors resulting from inaccurate reports.

### Model parameters

The Johns Hopkins University CSSE data repository (Johns Hopkins University Center for Systems Science and Engineering 2020) provides data on three time-series of Covid-19 indicators: daily counts of confirmed cases (*C*), recovered cases (*R*), and deaths (*D*). These data allow us to calculate additional time-series, such as:

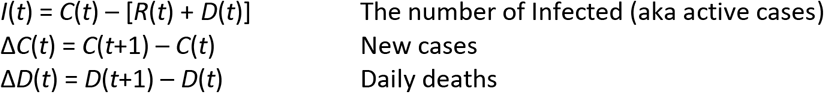

We use these data to calculate the daily estimates of the SIRD parameters, as described in the preceding section.

Next, we smooth the data using *ksmooth()*, a kernel regression function in R. We use the normal kernel and bandwidth of 10 for β and δ, and 20 for γ. Because data are very noisy when the total number of cases is still small, we smooth the data only after *C*(*t*) exceeds 100 cases and use a constant for the period before, set equal to the first smoothed estimate.

Using smoothed estimates of the model parameters we solve the SIRD model forward in time for the period of the epidemic. For initial values we assume *S*(0) = *N*, and *R*(0) = *D*(0) = 0. For *I*(0) we use a value that matches mean model-predicted *I*(*t*) to the number of infected in the data for the time period when the cumulative number of cases exceeds 100. In other words, *I*(0) is a parameter estimated from the data.

As a measure of model fit, we use the coefficient of prediction *R*^2^

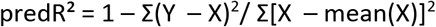

where *Y*(*t*) are model predictions (*Y* stands for *C*, *R*, etc), *X*(*t*) are data, and mean(X) is the mean of *X*. This measure, known as the coefficient of prediction, is similar to the coefficient of determination in regression. It also reaches a maximum of 1 for perfect prediction. Unlike regression *R*^2^, however, predR^2^ can be negative, when the model predicts data worse than the data mean. We calculate predR^2^ separately for log-transformed and not-log-transformed data.

Note that the purpose of numerically solving the SIRD equations is not to provide an independent test of the model. Using raw (unsmoothed) daily parameter values will generate a perfect “fit” of the model to data because these values were obtained from the data. Calculating trajectories with smoothed parameter curves, on the other hand, provides a check of internal coherence of the approach. In other words, the question becomes whether smoothing can still generate trajectories close to the data. The second reason to run the predicted trajectories is that initial counts typically severely underestimate the true number of infected. By chosing *I*(0) that fits the later course of the epidemic (after the number of infected exceeds 100) we obtain an estimate of the course of the epidemic before this threshold is reached. The difference between the predicted and observed *I* provides us with an informal estimate of the degree with which the initial numbers were underestimated.

### Case studies

In the rest of the document we report analysis results for each country (except for China, where we focus on the province of Hubei). Each case study begins with *Background*, which provides a narrative on the overall course of the Covid-19 epidemic in the focal country. This section is followed by *Analysis Results*, which are organized as folllows:

The top two rows show the data (points) and model predictions (curves) for (a) I, (b) D, (c) R, (d) ∆C, (e) ∆D, and (f) C.

(g) Transmission coefficient (β): daily estimates (points) and smoothed estimate (curve).

(h) Death rate (δ): daily estimates (points) and smoothed estimate (curve).

(i) Exponential growth rate of the epidemic (*r_t_*). The dotted line is *r_t_* = 0, above which the epidemic grows and below which it declines.

The final section, *Conclusions*, provides a brief summary of what has been learned by applying the SIRD model to the data for the country.

### Hubei, China

#### Background

On 31 December 2019, the WHO China Country Office was informed of a cluster of 27 pneumonia cases of unknown etiology that had emerged in Wuhan, the capital of Hubei province, China (World Health Organization, 2020a) (although an unverified report from the *South China Morning Post* suggested that a Covid-19 case traced back to 17 November 2019, in a 55-year-old from Hubei province, may have been patient zero (Ma 2020)). Cases within the cluster shared an epidemiological link with the Wuhan Huanan seafood wholesale market, which was shut down on 1 January 2020. At this stage, there was no clear evidence of human-to-human transmission (WHO 2020a). On 8 January 2020, results from sequencing analysis from lower respiratory tract samples revealed a novel coronavirus, which was then named “2019 novel coronavirus (2019-nCoV)” (F. Wu et al. 2020). On 11 January 2020, the first death from 2019-nCoV was reported in Wuhan (Bryson Taylor 2020). On 20 January 2020, Hubei reported 258 confirmed cases including six deaths (WHO 2020d) and by 29 January the virus had spread to almost all provinces of mainland China (7,736 confirmed and 12,167 suspected cases, 170 deaths) and to 18 countries around the world (82 confirmed cases) (WHO 2020g). By 8 February, over 723 deaths from the coronavirus infection had been reported in China and 34,598 were confirmed to be infected, of which 27,100 were in Hubei (WHO 2020k). As of 6 April 2020, the epidemic has led to 67,803 confirmed cases and 3,212 deaths in Hubei province.

The Chinese health authorities claimed to have promptly enforced intervention measures against the outbreak. These measures were implemented from 20 January 2020 and included intensive surveillance, epidemiological investigations of the cluster cases, identification and monitoring of contacts, and active case finding as well as risk communication to improve public awareness and promote self-protection measures (WHO 2020a). In a press conference on 20 January, the population was advised to avoid visiting Wuhan unless extremely urgent and to wear face masks. Moreover, the Chinese New Year celebrations were cancelled and health and temperature screening was implemented for travelers moving in and out of railway stations (X. W. Wu 2020). Commands for epidemic control (CEC) were set up in different regions, including Wuhan and Hubei. Many inter-province public transportation services were suspended. On 23 January 2020, the government of the People’s Republic of China imposed a lockdown in Wuhan and other cities in Hubei province. This was the first known instance in modern history of a major city with as many as 11 million people being locked down (Crossley 2020; Cyranoski and Silver 2020). The imposed lockdown included travel restrictions to and from Wuhan, a suspension of public transport within the city, and a mandatory home quarantine (citizens were only allowed to leave their home every two days, for a maximum of 30 minutes). Social distancing measures were enforced, including the cancellation of events and gatherings, closure of public places, schools and universities, and prohibition of outdoor activities (Lau et al. 2020; Zhu, Wei, and Niu 2020). On 23 January, the construction of an emergency field hospital (Huoshenshan Hospital) to treat patients infected with the coronavirus began, and was completed within 10 days. A second emergency hospital (Leishenshan Hospital) was built nearby and opened on 8 February (Holland and Lin 2020). New laboratories were also rapidly set up to increase testing capacity (Miller et al. 2020). Healthcare capacity was enhanced from 24 January onwards, with more than 30,000 medical workers sent to Hubei, two-thirds of which were sent to Wuhan (Ye 2020). On Wednesday 18 March, some travel restriction measures were lifted in Hubei.

#### Analysis results

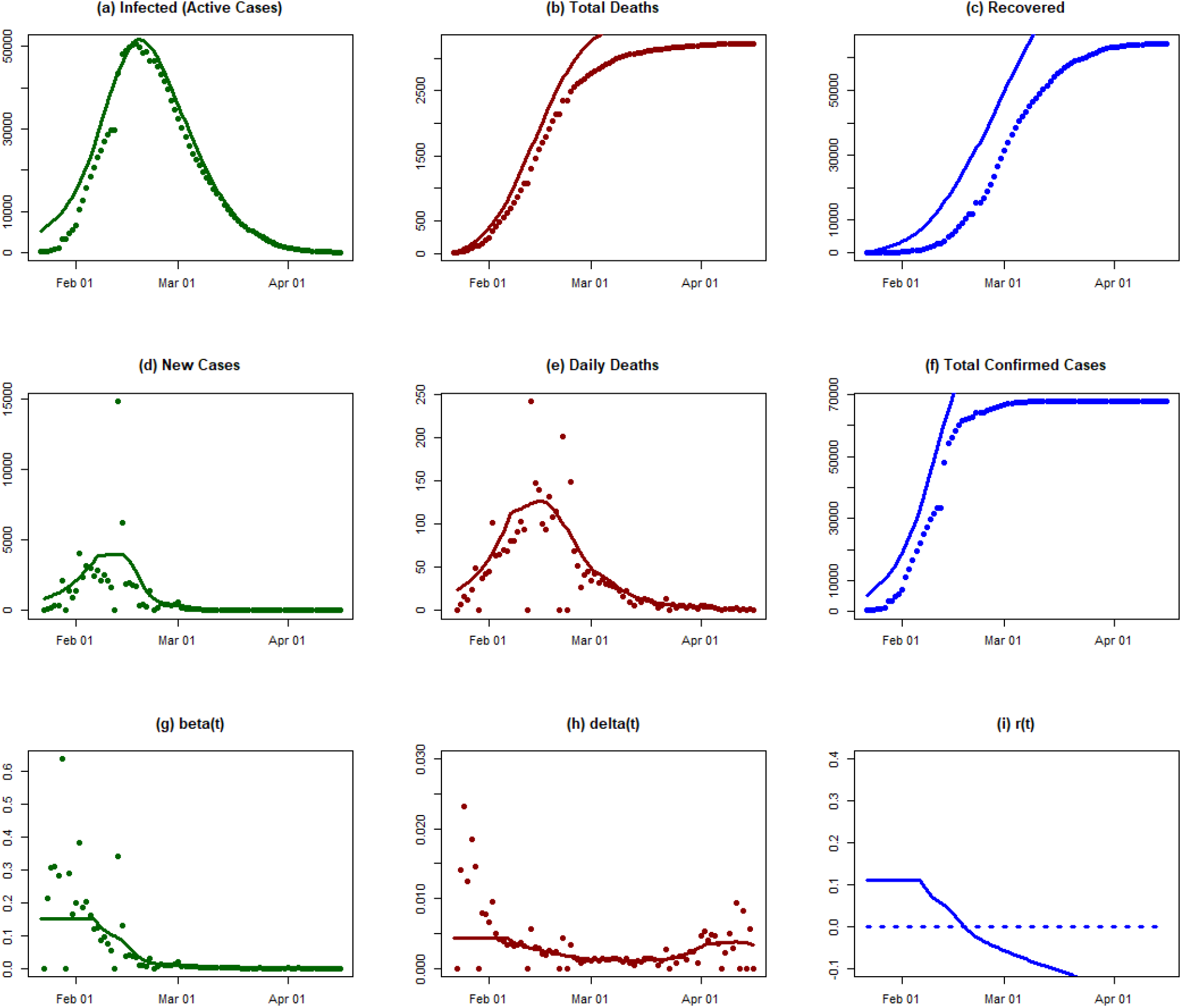

#### Conclusions

The dataset for China has some limitations. Indeed, the Chinese authorities revised the reporting of cases at least six times. On April 17 they suddenly accounted for additional >300 cases and >1000 deaths due to Covid-19. Since diagnostic dates for those additional cases are unknown, the results above were fitted to the data series ending on April 16. An overall poor fit between model-calculated trajectory and data suggests that the limitations in the data reporting mentioned above precludes us from obtaining an accurate understanding of the course of Covid-19 epidemic in Wuhan based on the data reported.

However, we note that Wang et al (2019) obtained a very good fit using a similar mode, which was fitted to data extracted from the Chinese municipal Notifiable Disease Report System (reference: https://doi.org/10.1101/2020.03.03.20030593). Below we reproduce a figure from this article. The implication from this comparison is that our failure to obtain a good fit was due to problems with the data we have available, rather than the course of epidemic in Wuhan deviating from the pattern predicted by the epidemiological model.

**Figure 4.**
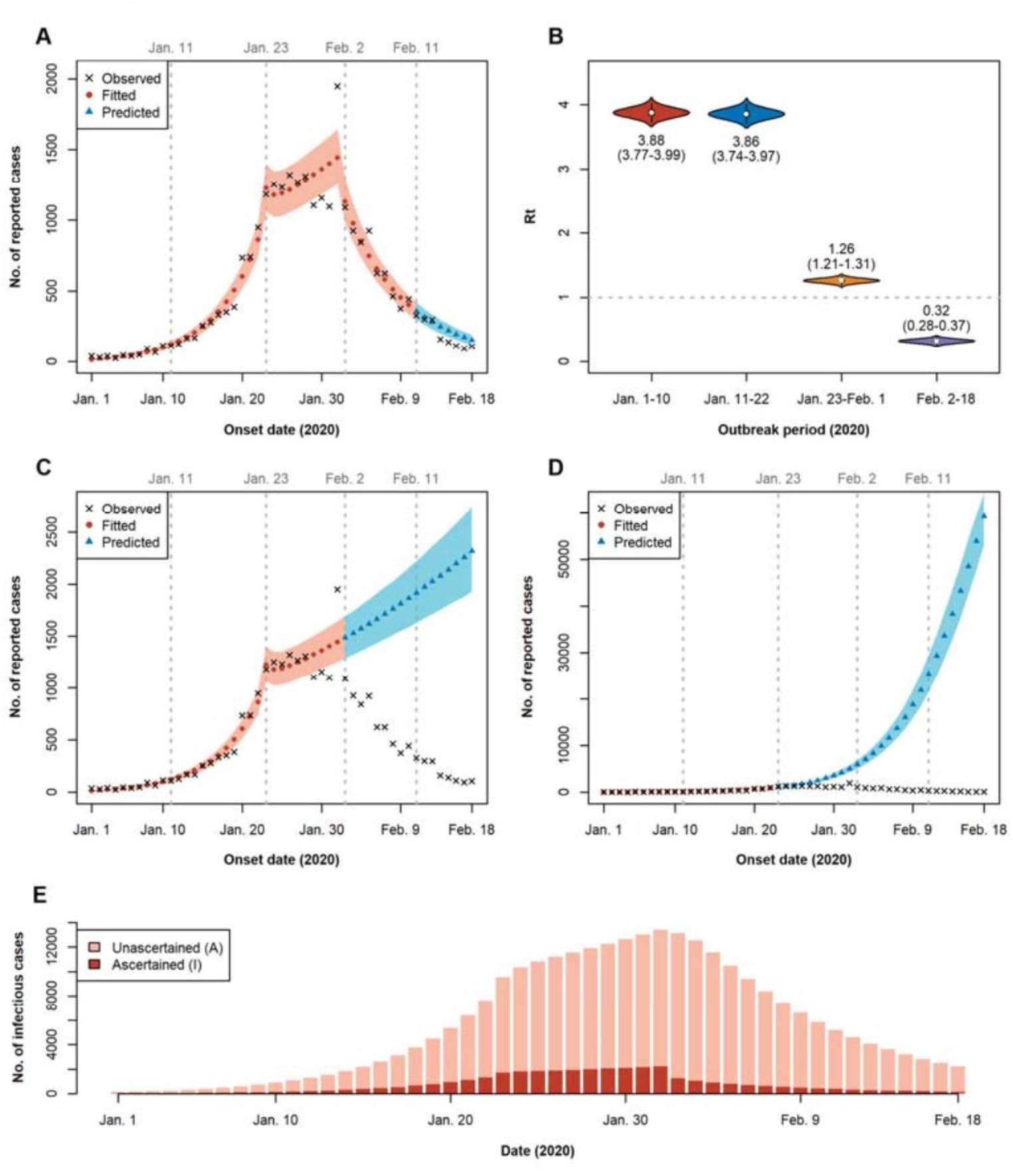
Four-period susceptible-exposed-infectious-recovered modeling of the Covid-19 epidemics in Wuhan.

### South Korea

#### Background

The first case of Covid-19 in South Korea was reported on 20 January 2020 and the number of cases increased in the following weeks (Duddu 2020; Shim et al. 2020). The number of confirmed cases increased significantly after 19 February (Johns Hopkins University Center for Systems Science and Engineering 2020; Shim et al. 2020). By 21 February, the cumulative number of infections had passed 100, with 155 confirmed cases and the first death from the virus. By a week later, on 28 February, there had been 2,022 cases and 13 deaths (Our World in Data 2020a; 2020d). This explosive growth in infections was linked to “Patient 31,” a member of the ShincheonjiChurch of Jesus in the city of Daegu who has been described as a “superspreader” (Gregory 2020; Shin and Cha 2020). However, measures taken by the government to contain the Daegu cluster and control the spread of the virus have been successful. On 15 March, the total number of cases stood at 8,162 and deaths at 75 (Our World in Data 2020a; 2020d). As of 2 April, there have been 9,976 confirmed cases, of which 169 have died and 5,828 have been reported as recovered (Johns Hopkins University Center for Systems Science and Engineering 2020). South Korea has therefore achieved an unusually low case fatality rate so far, keeping its Covid-19 death toll below 300.

The government response to the first cases of Covid-19 in the country focused on contact-tracing and high-tech surveillance through smartphone data, closed-circuit television footage and credit card data (Sonn 2020).

Mass testing has been a cornerstone of South Korea’s approach. By mid-March around 20,000 people—including those without symptoms—were being tested per day (Bicker 2020; Edward 2020). Drive-through testing centres have been used to protect healthcare workers and reduce delays caused by the need to disinfect hospital facilities after each patient. Both those who test positive and their contacts within the past 14 days were quarantined and smartphone apps were used to monitor their movements (Kasulis 2020). As of 1 April, 421,547 people have been tested (Statista 2020).

Following the identification of the Daegu cluster, emergency measures were put in place to contain the outbreak: extra testing centres, contact-tracing and monitoring of Shincheonji members in Daegu and beyond, and case isolation (Kim 2020; Korean Centers for Disease Control and Prevention 2020). No national lockdown was implemented: many restaurants and other businesses stayed open but checked customers’ temperature before admitting them (Choe 2020).

The government has also sought to encourage social distancing and respiratory etiquette among the population. Schools were closed in late February and as of 2 April they have not yet reopened (BBC News 2020f; Ock 2020; Shin 2020). Many policies have relied on voluntary public cooperation. Large-scale public information campaigns promoted wearing masks, washing hands often and thoroughly, and maintaining social distance (Chunghoon and Soh 2020).

#### Analysis results

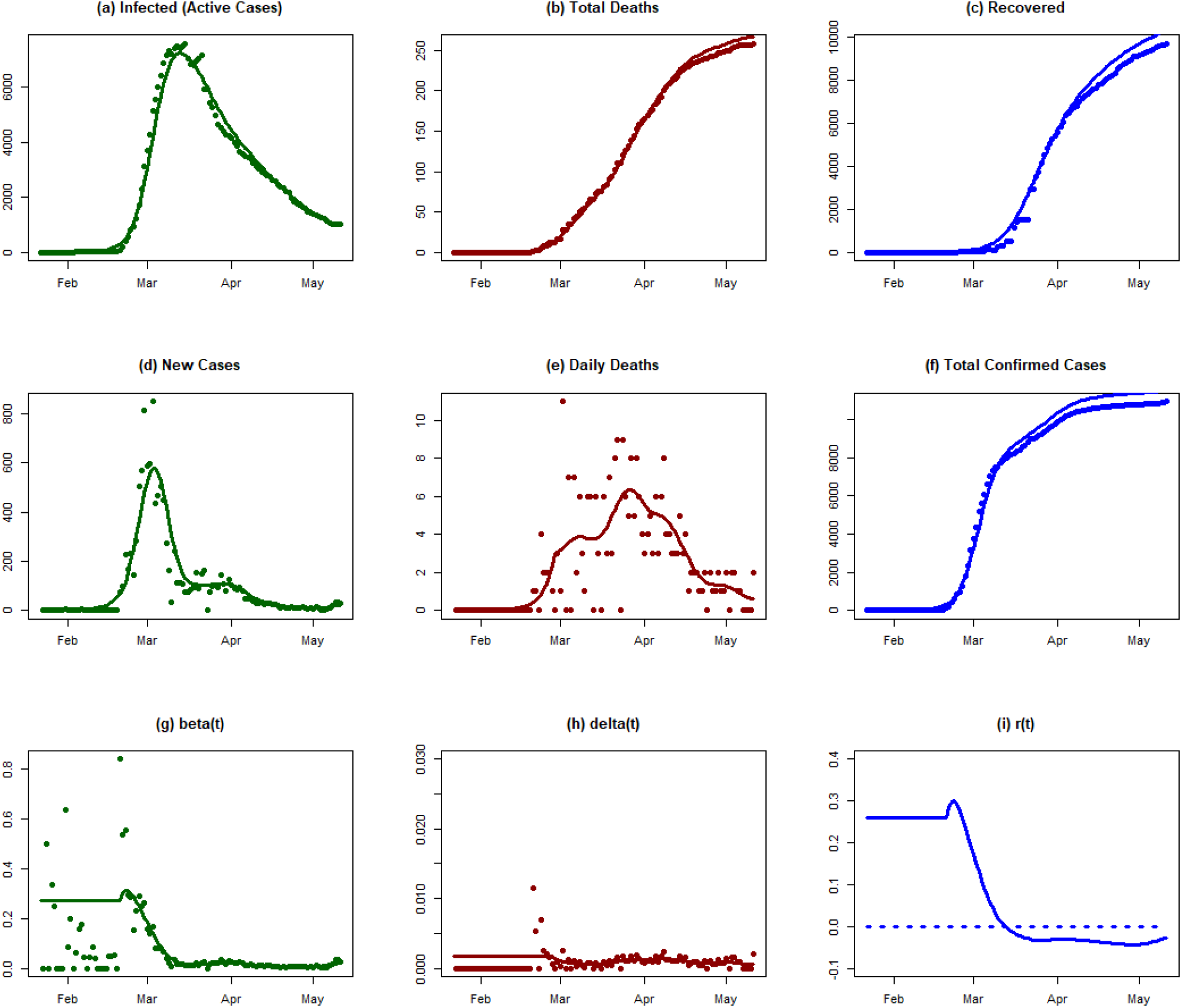

#### Conclusions

Although South Korea did not enforce a national lockdown, the measures implemented by the government were very effective in stopping the disease. The transmission rate declined very rapidly within a ten-day period centered on 1 March. This caused the exponential rate of disease to cross the zero-line around 10 March. Currently, all signs indicate that the epidemic is under control (ref).

### Japan

#### Background

On 15 January 2020, the first case of Covid-19 was confirmed in Japan in an adult male, with a history of travel to Wuhan, China (WHO 2020c; 2020b). Up to 1 February 2020, 17 confirmed cases had been reported to the WHO (WHO 2020h). The first death due to Covid-19 in Japan was confirmed on 14 February 2020, while 33 cases were confirmed (WHO 2020m). On 1 March 2020, there were 239 confirmed cases and 5 deaths (WHO 2020p). On 1 April, the total number of confirmed cases was 2,178 with 57 deaths (WHO 2020z). Four days later the number of new confirmed cases had increased by 66%, counting 3,271 cases and 70 deaths (WHO 2020ab). Approximately one-third of these confirmed cases were from the Tokyo region (Knüsel 2020). At the end of March, the new Covid-19 infections could no longer be traced back to known sources, bars, karaoke clubs and other entertainment venues being the most probable places of infection (Knüsel 2020). Some schools in Japan, in municipalities with no Covid-19 cases, are set to reopen in April (Siripala 2020). However, many public schools in Tokyo will remain closed until early May. In the second week of May, a month-long state of emergency for the Japanese regions of Tokyo, Kanagawa, Saitama, Chiba, Osaka, Hyogo and Fukuoka was declared (Johnston 2020; McCurry 2020). The state of emergency creates the opportunity to implement at subnational levels (prefectures and municipalities) more effective and specific measures to mitigate the spread of Covid-19 (Johnston 2020; McCurry 2020).

As of 3 April 2020, a total of 39,446 people in Japan had undergone PCR tests for COVID-19 (Statista Research Department 2020) (0.00031% of the total population).

#### Analysis results

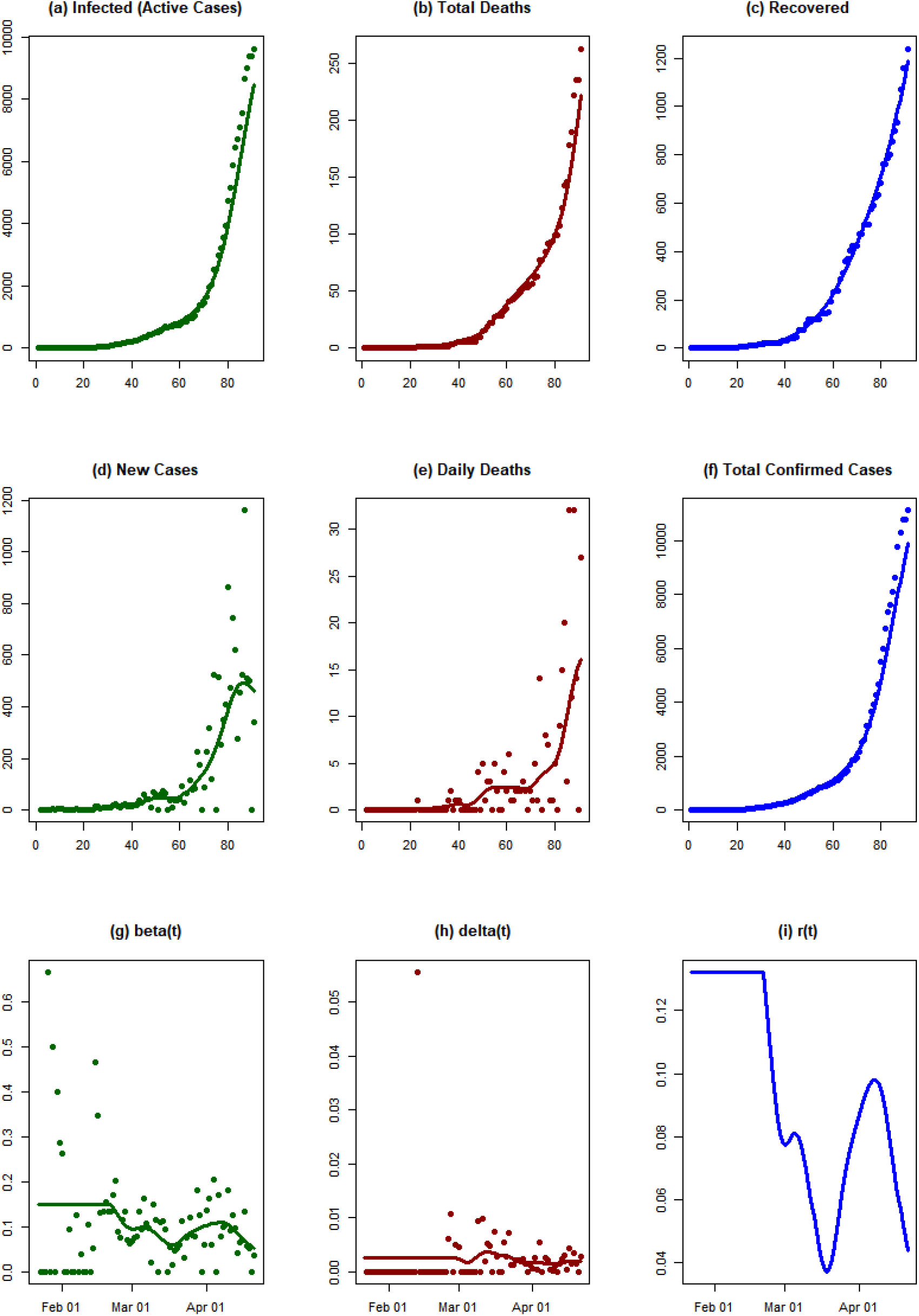

#### Conclusions

The number of cases was low initially in Japan. However, we observe an explosive growth in the number of infected people around the end of March and the beginning of April 2020. As of 21 April, Japan was able to reverse the exponential growth rate of epidemic, but it has not crossed the zero line yet. The decay in exponential growth rate might be related to the strict measures taken by the Japanese government, such as a state of emergency in seven prefectures on 7 April, which was further extended to the whole country on 16 April.

### India

#### Background

On 30 January 2020, India reported its first case of Covid-19 to the WHO (WHO 2020g; Reid 2020). In total, three confirmed cases were reported by 1 March 2020 (WHO 2020p), but by 1 April, the number of confirmed cases had increased to 1,636, with 38 deaths (WHO 2020z). In January, the Indian government announced a travel alert for Indian citizens, in particular for the city of Wuhan (Yan and Wallen 2020). Screening of people at airports began on 17 January and approximately 600,000 passengers had been screened for symptoms up to 3 March (Biswas 2020). Between 9 and 23 March, more and more Indian federal states closed their land borders with neighbouring countries, followed by a travel ban for non-neighboring states, e.g. the European Union. Other countries, such as Nepal and Bhutan, closed their borders with India (*The Hindu* 2020; Laskar 2020; ET Bureau 2020). All Indian states were required to implement social distancing measures from 16 March (Sharma 2020).

On 24 March, the Indian government announced a national lockdown for the next 21 days (BBC News 2020g), although some Indian states have extended the lockdown until 30 April (Hindustan Times 2020). In this context, universities, schools, all transports and services have been suspended (exception e.g. essential goods, emergency services, supermarkets and hospitals). Testing coverage was increased from 20 March: people with symptoms suggesting possible Covid-19 infection, particularly those of pneumonia, were tested regardless of travel or contact history (Ghosh 2020). As of 8 April, a total of 127,919 samples have been tested (Findlay 2020) from which 5,194 were positive, and 149 deaths have been reported to the WHO (WHO 2020ac).

#### Analysis results

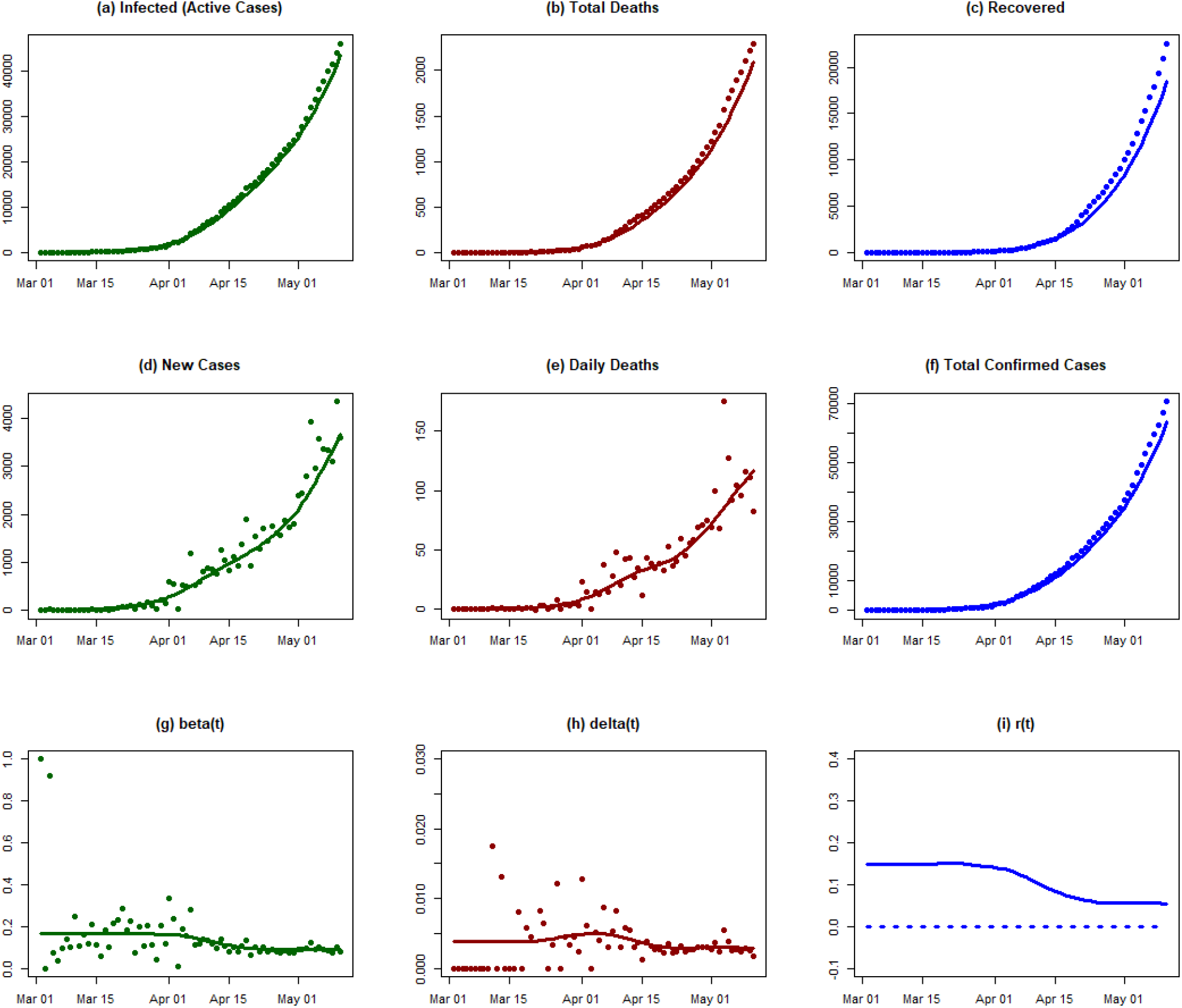

#### Conclusions

We observe a growth in the number of infected individuals between the middle of March and the first week of April 2020 in India. As of 11 May, the exponential growth rate of epidemic has decreased, but it has not crossed the zero line yet. The decay in the exponential growth rate of the epidemic could be related to various measures, including the enforcement of a national lockdown. What is worrying is that the growth rate of the epidemic stopped declining in mid-April. Thus, as of 11 May, the government has not succeeded in controlling the epidemic.

### Austria

#### Background

On 25 February 2020, the first two confirmed cases of Covid-19 in Austria were reported, in Innsbruck. Both patients had a travel history to Bergamo, Italy (ORF.at 2020a). Both were admitted to hospital and were later discharged, on 5 March (ORF.at 2020b). All contact persons were traced and tested (Al Jazeera 2020a). By 1 March, 1,826 tests had been conducted (Tiroler Tageszeitung 2020). The first Covid-19 death in Austria was reported on 12 March; the patient had a recent travel history to Italy (ORF.at 2020c; WHO 2020s).[6-7]. One month later (on 26 March), there were 5,888 confirmed cases and 34 deaths (WHO 2020w). On 3 and 4 April 2020, there were more newly recovered than newly infected people (on 3 April: 11,383 infected and 2,022 recovered and on 4 April: 11,665 infected and 2,507 recovered (Bundesministerium für Soziales, Gesundheit, Pflege und Konsumentenschutz 2020a; 2020b)).

On 10 March, the first universities closed, airport and health checks (especially at the border crossings to Italy (Anschober 2020) were implemented, and people returning from Italy were quarantined. On 10 March, mass gatherings were prohibited (i.e. of over 100 people indoors or over 500 outdoors) (Kurier 2020). All schools were closed from 16 March (Peitler-Hasewend and Jungwirth 2020), as of 8 May 2020 they remain closed but are scheduled to reopen in mid-May 2020 (ORF.at 2020j).

On 13 March, a cordon sanitaire was implemented around the municipalities of Ischgl, Kappl, See, Galtür and St. Anton am Arlberg in Tyrol (ORF.at 2020e) and four days later (17 March) around the entire federal state of Tyrol (Van der Bellen and Kurz 2020).[13] On 7 April, the cordons sanitaires were lifted (except for St. Anton, the Paznaun valley and Sölden) (Kleine Zeitung 2020a). On 14-15 March the Austrian parliament passed the Covid-19 Law, ordering the closure of all non-essential shops from 16 March (ORF.at 2020d). Initially bars, restaurants and cafes were allowed to open until 3 p.m. (Kleine Zeitung 2020a), but from 17 March all restaurants and bars were closed completely (ORF.at 2020f). With the Covid-19 Law (ORF.at 2020g), far-reaching restrictions on individual movement were imposed. Social distancing measures were implemented, e.g. direct patient contact with doctors was reduced by issuing electronic prescriptions over the phone. The law also required people to keep a distance of 1–2 meters from others and banned gatherings of more than five people. On 20 March, further social distancing measures were introduced: children’s playgrounds and sports grounds were closed and working from home was encouraged. On 25 March, the “Stopp Corona” tracking app was released, which allows users to be notified when they have come into contact with a person infected with Covid-19 (Österreichisches Rotes Kreuz 2020).

From 6 April, it became compulsory to wear a basic face mask, covering the mouth and nose, inside supermarkets, and from 14 April this requirement was extended to public transport (ORF.at 2020i; 2020h). Beginning on 14 April, small shops under 400 m^2^, hardware stores and garden centres were allowed to reopen with adaptative measures, i.e. no more than one customer per 20 square meters. From 1 May, other shops and hairdressers will also be able to reopen in Austria (Der Standard 2020).

#### Analysis results

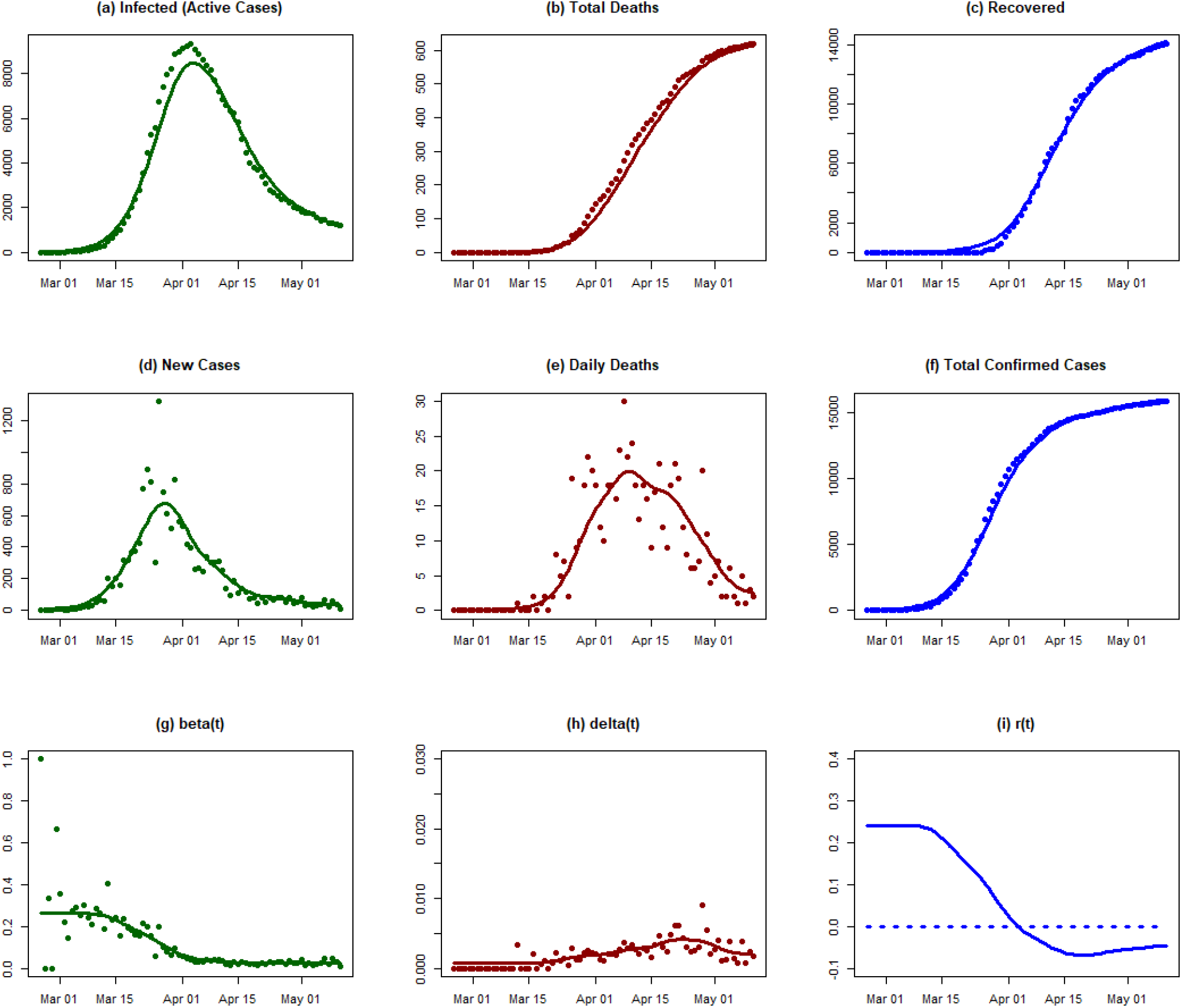

#### Conclusions

Austria gradually shut down in early March and by 15 March the implemented measure may have decreased the transmission coefficient. The progress was rapid and by early April the exponential growth rate of the epidemic crossed the zero line. Under the current conditions the epidemic is showing clear signs of dying out.

### USA

#### Background

The first confirmed case in the United States was reported to the WHO on 23 January 2020 (WHO 2020e). On 1 March 62 cases had been confirmed (WHO 2020p). On 3 May, a total of 1,093,880 confirmed cases and 62,406 deaths had been reported (WHO 2020ah).

The United States has introduced a series of travel restrictions. Foreign nationals with a travel history within the past 14 days to specific countries, such as China (from 2 February) and Iran (2 March), were barred from entering the country, while Americans returning from these countries were ordered to self-quarantine for 14 days (University of Colorado Boulder 2020; Robertson 2020). On 13 March a national state of emergency was declared (Liptak 2020a). On 16 March the government advised against meetings of over 10 people (Liptak 2020b).

New York has been particularly severely affected by the epidemic, the state of emergency was declared on 7 March. Several executive orders requiring progressively more non-essential workers to stay at home were issued. All mass gatherings of over 500 people were cancelled from 13 March, and events with fewer than 500 attendees were ordered to “cut capacity by 50 percent” (The New York State Governor 2020). Effective from 22 March, the “New York State on PAUSE” executive order ordered “all non-essential businesses statewide” to close (NY State Department of Health n.d.). This order also directed the population to suspend all non-essential social contact, maintain a distance of six feet from other people in public, limit the use of public transport to “when absolutely necessary,” and to self-isolate at home if they became sick.

Ohio’s state government closed all bars and restaurants on 15 March, and was followed by Illinois (Conradis 2020), New York, New Jersey, and Maryland (Education Week 2020). On 16 March, the national government announced that citizens should stay home and avoid contact for 15 days, and these social distancing measures were then extended until the end of April (Keith et al. 2020). Lockdowns were implemented at different times in different states, such as in Idaho on 25 March and Alabama on 4 April (Education Week 2020). The majority of schools nationwide have been closed since 10 April (Education Week 2020). At the beginning of April, the governments of several states and cities, including New York, Los Angeles, Colorado, and Pennsylvania, have asked citizens to wear face masks in public spaces (Education Week 2020; Durkin 2020; Briggs, Wolfman-Arent, and MacDonald 2020).

#### Analysis results

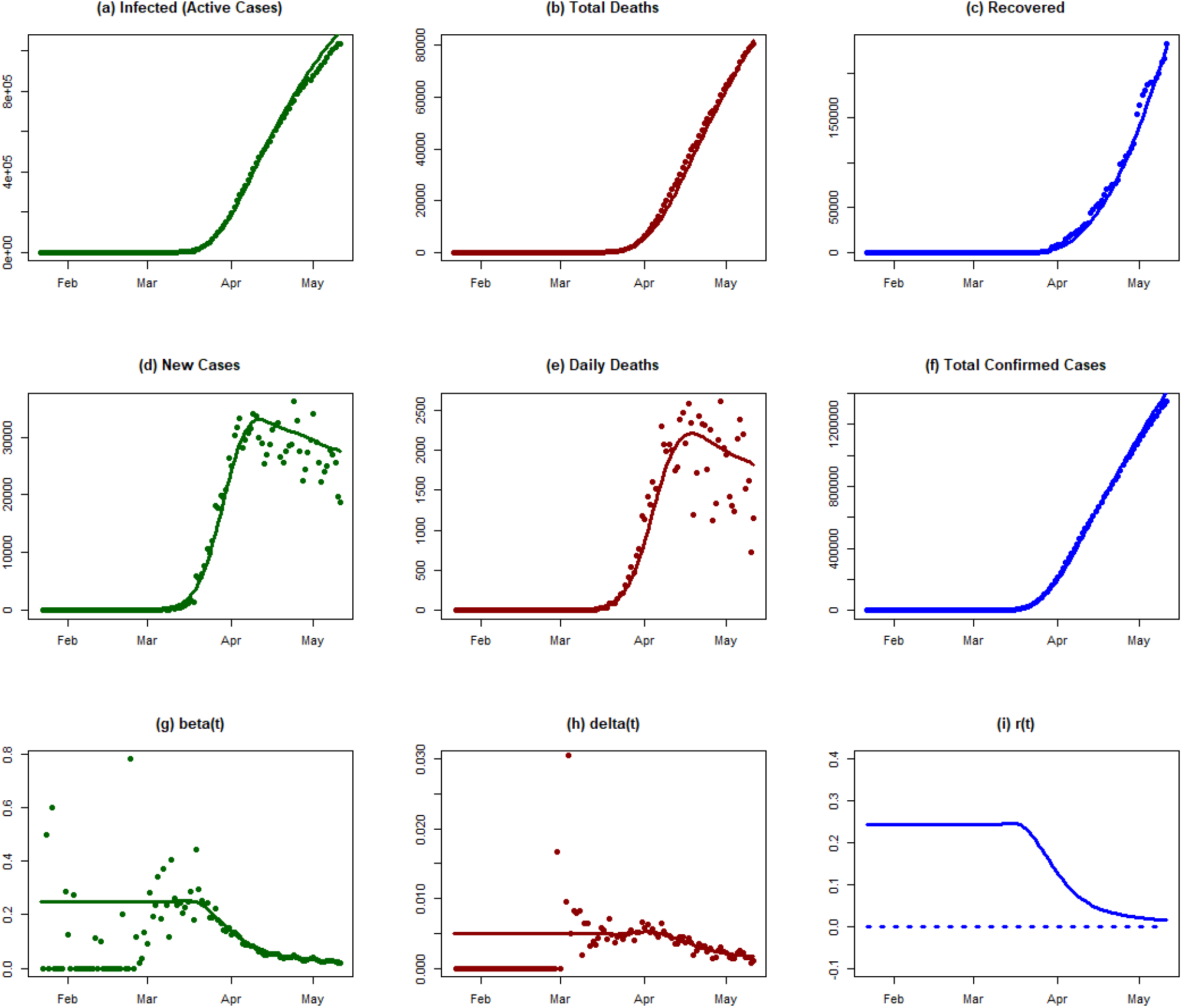

#### Conclusions

The early course of the Covid-19 epidemic in the USA (before March) cannot be resolved with the present data. The reporting rate clearly improved after 1 March. For the first three weeks in March, the epidemic grew nearly exponentially with a transmission rate of nearly 0.25 per day. Beta started to decline only towards the end of March. Unfortunately, by 11 May beta was still at 0.025, which was enough to slow down the growth of epidemic but not enough to bring it to an end. As of 11 May, the exponential growth parameter, r(t), is still positive.

### Taiwan

#### Background

The first confirmed case in Taiwan was reported on 21 January, and according to the Central Epidemic Command Center (CECC), set up on 22 January, 124 separate measures have been implemented since the beginning of the pandemic (Wang, Ng, and Brook 2020). Taiwan began to respond to the pandemic early on, before the first confirmed cases. The government introduced inspection measures for all incoming flights from Wuhan, China from 31 December 2019. Health screening was implemented for all incoming travellers from Wuhan on the same date (Wang, Ng, and Brook 2020).

Contact tracing started on 5 January: the government started monitoring travellers who had been to Wuhan, China in the past 14 days. Individuals classified as high-risk according to their recent travel history were ordered to self-quarantine (Wang, Ng, and Brook 2020). On 14 February the government launched the Entry Quarantine System for contact tracing, an online health declaration platform for all incoming travellers upon arrival (Wang, Ng, and Brook 2020).

The government promoted the wearing of face masks as a preventative measure and banned the export of face masks on 24 January. To prevent citizens from hoarding face masks and raising the price, the government implemented a mask rationing system, which limited the quantity each citizen could buy and when they could buy them, based on their health insurance number (Ku and Kao 2020). The government increased face mask production to 8 million per day and formed 60 new production lines to meet the demand (Yang 2020). By 7 April, daily testing capacity had increased to 3,800 (Taiwan Centers for Disease Control 2020b).

Travel tours from China were suspended in February, and this restriction was subsequently extended to other high-risk regions. The number of confirmed cases rose rapidly in March due to the high volume of incoming travelers (The Straits Times 2020). The government tightened its airport and border control by denying entry to foreign nationals from 19 March and imposing a 14-day self-quarantine for all incoming travelers (Chang, Huang, and Chen 2020). Social distancing measures have also been implemented, including the cancellation of mass gatherings of over 100 people on 25 March (Taiwan Centers for Disease Control 2020c; 2020a).

#### Analysis results

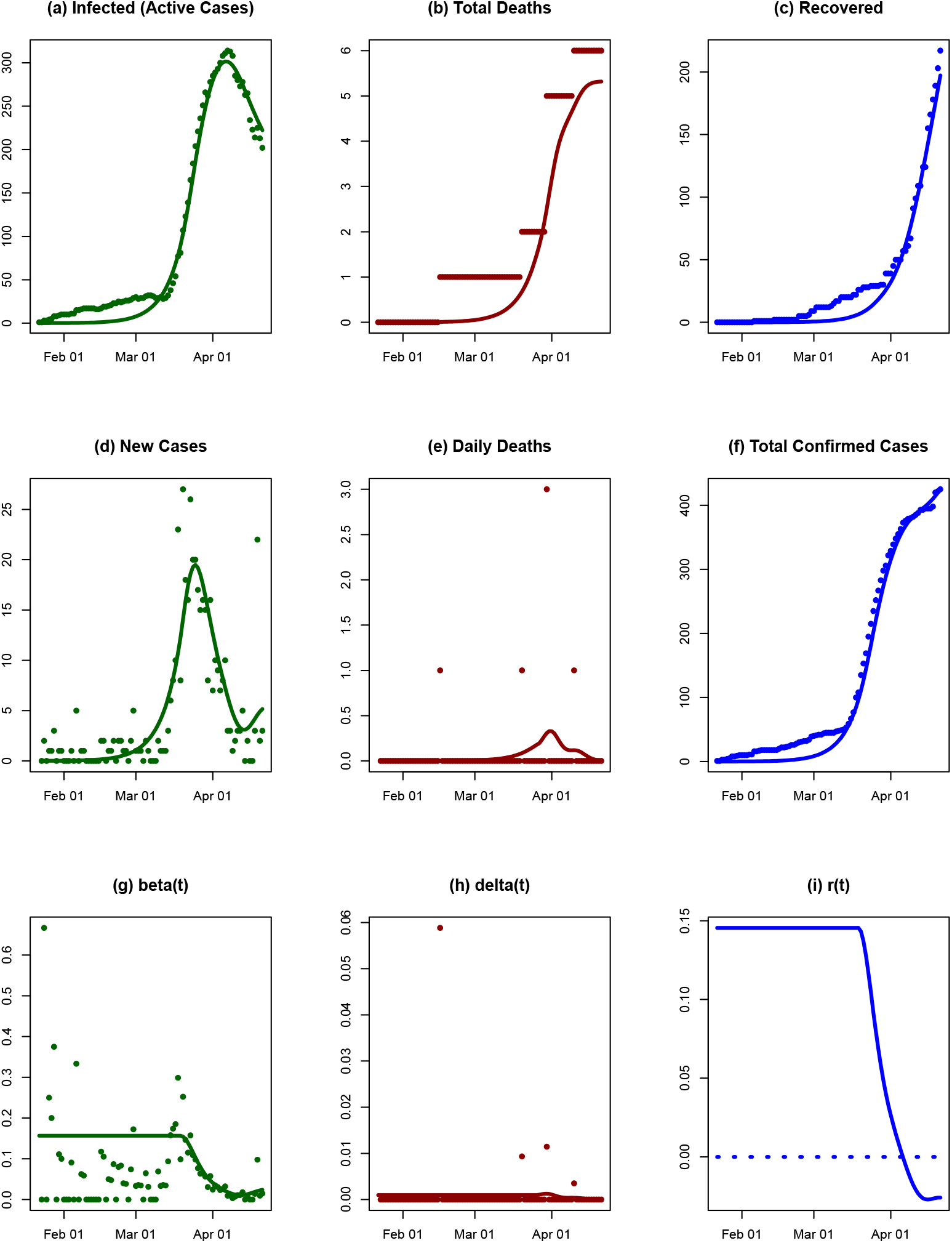

#### Conclusions

The time series runs from 22 January to 21 April 2020, with confirmed cases ranging from 1 to 422. The relatively low active cases and infection rate may be due to the Taiwanese government’s early interventions: case identification and health checks at airports from 31 December 2019, travel restrictions denying entry to people who had been in Wuhan from 22 January 2020, and social distancing measures (event cancellations and closure of educational institutions) in February 2020. Additional quarantine measures were introduced on 19 March 2020.The government has increased testing capacity since 24 January 2020 (when there were just three confirmed cases), which is an important measure for targeting the source of the spread.

The overall trend of the R(t) and the SIRD model suggests that the infection rate and number of active cases rose rapidly after day 40. The SIRD model-fitting did not perform too well using data from Taiwan, with a mean R2 of 0.76. In particular, the daily death rate was estimated poorly. The confirmed cases were underestimated prior to day 40. Taiwan implemented a series of measures early on during the pandemic, which may be the reason for the low number of cumulative active cases, and relatively low R(t) until 8 March. The surge in the number of cases was mostly due to imported cases, which cannot be addressed by one or two intervention measures alone. The transmission rate in the country decreased around mid-March. The effectiveness of increased testing capacity and contact-tracing measures may be reflected in the rising infected cases from day 58. The exponential growth rate of the epidemic decreased rapidly from around the end of March to mid-April, and the growth rate has crossed the zero-line. The fitted model suggests that the epidemic is slowing down in the country and that the interventions were effective.

### Australia

#### Background

The first confirmed case in Australia was reported on 25 January 2020 (Australian Government Department of Health 2020), and as of 13 April there were 6,322 confirmed cases (Our World in Data 2020b). The government barred entry to travellers from China on 1 February as the virus spread rapidly there, and subsequently imposed an additional travel ban from other high-risk countries, such as Italy and South Korea (Bagshaw 2020; Bagshaw and McCauley 2020; Pannett 2020). Prime Minister Scott Morrison declared a human biosecurity emergency on 18 March, which allowed the government to enforce restrictions on individual movement and regional lockdowns (Pandey 2020). On the following day, Australia closed its border to all foreign nationals not resident in the country (Brockett and Scott 2020). On 22 March it was announced that no more than one person per four square metres would be allowed in indoor public places such as bars and restaurants, while other social distancing measures (including closing educational institutions and non-essential services) varied from county to county (Boseley and Knaus 2020). Mass gatherings of more than 500 people were suspended on 13 March, and more stringent measures limiting gatherings to two people came into force on 29 March (ABC News 2020; Worthington 2020). In addition to travel restrictions and social distancing, measures designed to prepare the healthcare system have been introduced, such as increasing the number of ventilators available and enhancing testing capacity (Lawson 2020; Tadros, McIlroy, and Margo 2020).

#### Analysis results

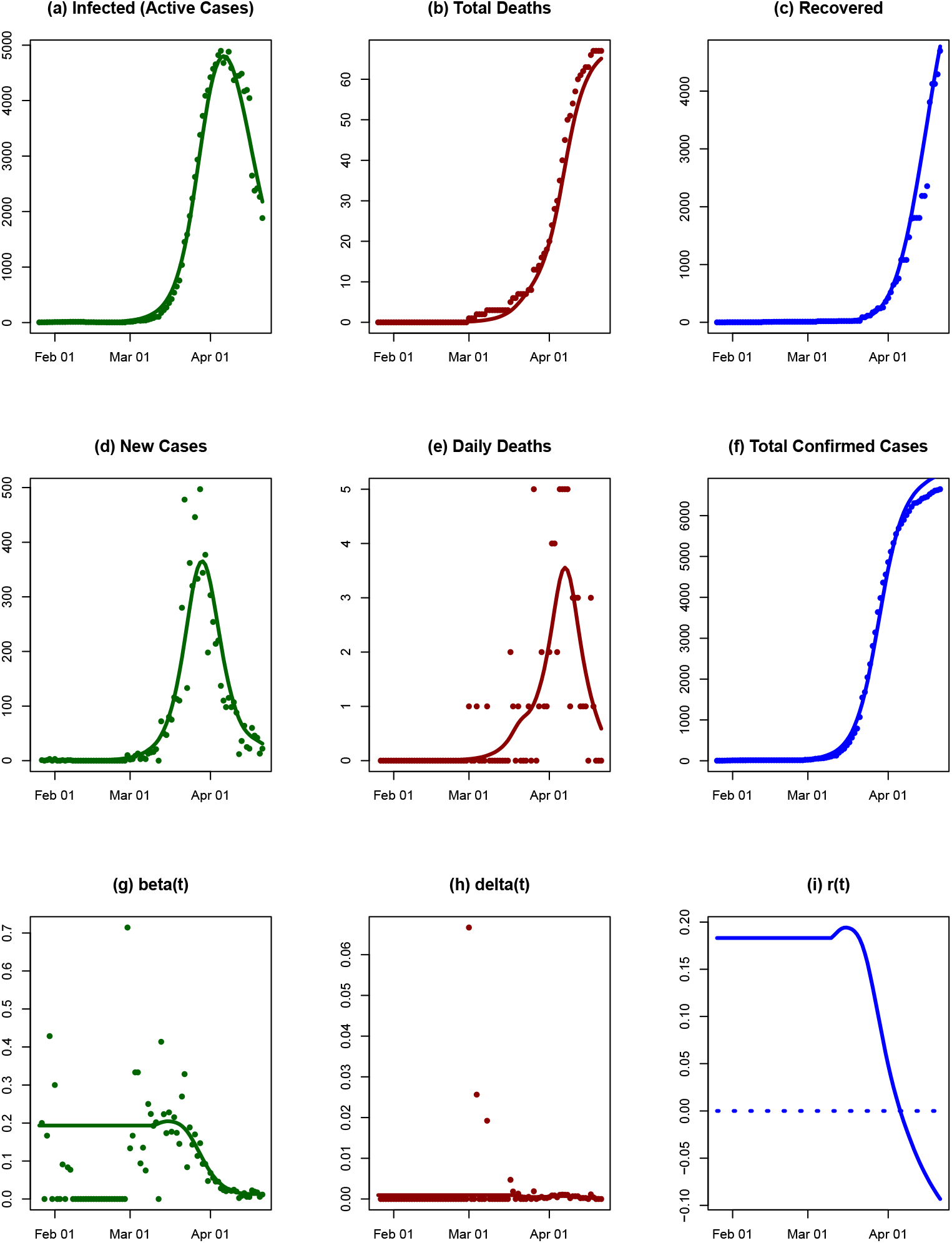

#### Conclusions

The model fitted with mean R2=0.91. The infected cases reached a peak around day 70, and the number of active cases began to decrease after this point. The total deaths showed similar a pattern to the new cases. The death rate was very low and flat during the observation period, and the transmission rate has been decreasing from the beginning of March. There was a growth in the number of infected cases around 15 March, which may due to the increasing testing capacity. Subsequently the growth rate of the epidemic declined rapidly and crossed the zero-line. The fitted model showed signs that the epidemic is slowing down and is under control in the country.

### Italy

#### Background

On 20 February 2020, the first case of Covid-19 was notified in Italy, in the Lombardy region. In the following week, the number of cases increased rapidly in the southern part of Lombardy (Cereda et al. 2020). On 22 February 2020, 60 additional cases as well as the first two deaths were notified (Dong, Du, and Gardner 2020). As the epidemic progressed, it spread to other Italian regions. On 19 March 2020, the number of deaths due to Covid-19 in Italy surpassed that of China. As of 7 April 2020, Italy is one of the biggest active hotspots of Covid-19 worldwide, with a total of 132,547 confirmed cases and 16,523 deaths (Dong, Du, and Gardner 2020). However, the true number of cases has probably been underestimated due to limited testing capacity (Flaxman et al. 2020).

On 31 January, the Italian government declared a state of emergency and all flights to and from China were suspended. On 21 February a cordon sanitaire was imposed around 10 municipalities in northern Italy (Desvars-Larrive et al. 2020). The cordon sanitaire around the epicentre of the outbreak in Codogno appears to have played a critical role in controlling the infection (Cereda et al. 2020). On 2 March 2020, as Italy reached 100 deaths, the government announced the complete closure of all educational institutions. On 9 March 2020, a national lockdown was imposed (Desvars-Larrive et al. 2020). Two days later, the government ordered the closure of all bars, restaurants and non-essential shops (Sylvers and Legorano 2020). On 19 March 2020 non-essential individual movements were prohibited, including all outdoor activities, and public places closed down. On 23 March 2020, all non-essential companies and industries were closed (Desvars-Larrive et al. 2020).

#### Analysis results

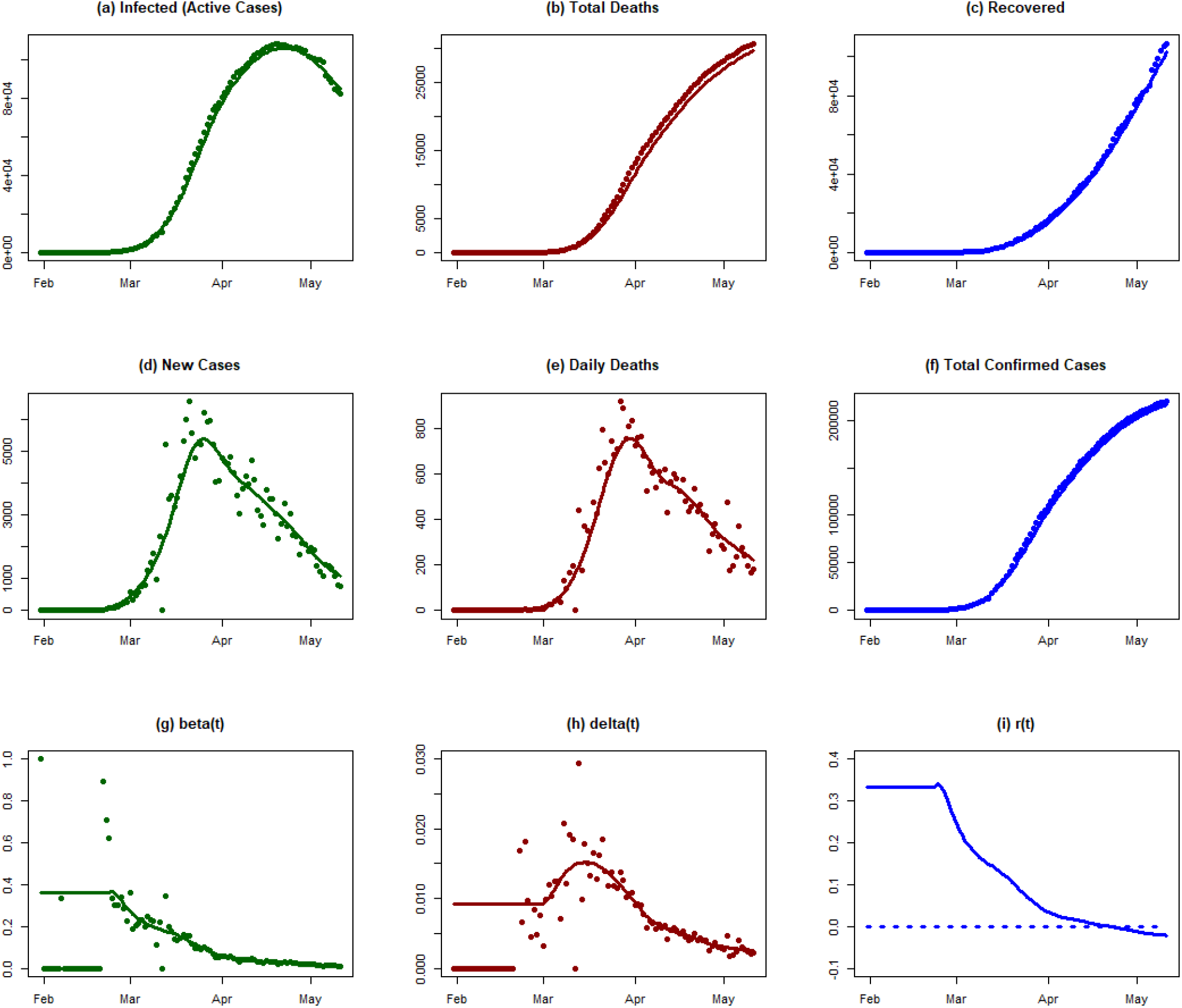

#### Conclusions

Although Italy has declared a state of emergency on 31 January, effective measures to slow down the rate of spread of the epidemic required more than a month to take effect. The transmission rate began declining after 1 March. Meanwhile, the death rate increased, reaching a peak in mid-March, probably because the healthcare system of Italy was overwhelmed by the number of patients requiring intensive care and respirators. The measures taken by the government finally bore fruit when the exponential growth rate of the epidemic crossed the zero line in late April.

### Spain

#### Background

Spain has been one of the countries worst-hit by the pandemic. The first few cases of Covid-19 were identified outside peninsular Spain and were linked to tourism in the Canary Islands. On 31 January, a German tourist on the island of La Gomera tested positive (Linde 2020). In late February a doctor from Lombardy (northern Italy) on holiday in Tenerife was found to have contracted the virus, causing the hotel he had been staying in to be locked down for two weeks (Jones, Boseley, and Burgen 2020; BBC News 2020c). Cases on the Spanish mainland began to take off in early March. The number of confirmed cases for the whole country by 29 February was 34; this had risen to 5,753 just two weeks later (15 March), and by 31 March 78,797 people had tested positive (Our World in Data 2020c). Spain overtook China in the number of confirmed cases on 30 March (Laudette and Landauro 2020). With 140,511 cases as of 7 April, the country is now second only to the USA in the scale of its epidemic (Johns Hopkins University Center for Systems Science and Engineering 2020). Its death rate is also very high: again as of 7 April, 13,897 people have died and 43,208 have been recorded as recovered (Johns Hopkins University Center for Systems Science and Engineering 2020).

Both the Spanish government and the governments of the autonomous communities have been criticized in the international press for a delayed response to the pandemic, despite the crisis unfolding in nearby Italy (Tremlett 2020; Ward 2020). An Interministerial Coordination Committee (Comité de Coordinación Interministerial) was created on 4 February to spearhead the country’s response (Ministerio de Sanidad, Consumo y Bienestar Social 2020; Desvars-Larrive et al. 2020). However, testing was initially slow to take off, and medical professionals warned that they lacked the resources needed to test all those showing symptoms (Güell, Sevillano, and Linde 2020).

Border restrictions were introduced gradually: flights between Spain and Italy were suspended on 11 March (Ministerio de la Presidencia, Relaciones con las Cortes y Memoria Democrática 2020), and on 16 March Spain announced that it would not allow foreign nationals to enter through its land borders except in special cases (Al Jazeera 2020c).

An important turning point in Spain’s approach was the declaration of a state of alarm on 14 March. Before that point, some social distancing measures had been implemented, such as the closure of schools by all autonomous communities between 9 and 12 March (Mateo and Ferrero 2020; RTVE.es 2020) and a ban on events with over 1,000 people by the Catalan government on 11 March (Rodríguez 2020). But on 8 March, “sports events, political party conferences and massive demonstrations to mark International Women’s Day all took place” (Tremlett 2020). The state of alarm signaled a much more restrictive approach, effectively a complete lockdown: citizens were ordered to stay at home unless going to work, shopping for necessities or helping others; cafes, restaurants and entertainment venues were closed (Zafra, Galocha, and Alonso 2020). Even leaving the house for exercise (such as a short walk, jog or bike ride) was banned. On 29 March the lockdown was tightened: all non-essential workers were ordered to stay at home (Marcos 2020). Prime Minister Pedro Sánchez announced on 4 April that the state of alarm would be extended until the 26th of the month (Cué, Junquera, and Vizoso 2020).

#### Analysis results

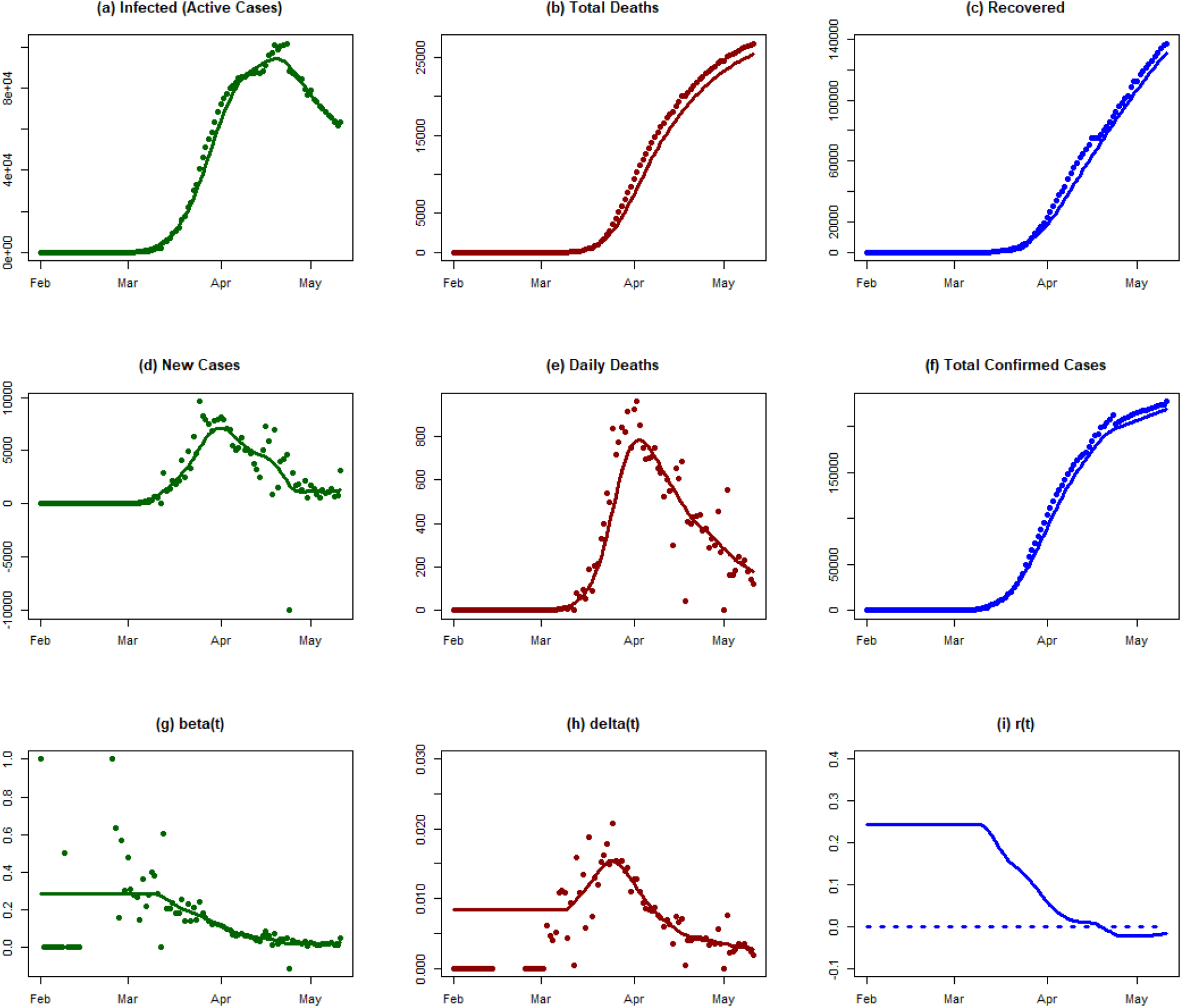

#### Conclusions

Spain is another example in which lockdown was implemented slowly over an extended period of time. As a result, the transmission rate began declining only in mid-March. What’s worse, the death rate increased substantially in the second half of March, perhaps because the hospitals were overwhelmed. The exponential growth rate has been declining, and finally crossed the zero line in late April.

### Turkey

#### Background

Turkey announced its first case of Covid-19 relatively late in comparison with nearby countries, on 11 March 2020 (McKernan 2020; Our World in Data 2020e). By a week later (19 March), the confirmed cases had jumped to 191 and the first death had been recorded. By the end of March, 9,217 cases and 168 deaths had been officially recognized according to the European Centre for Disease Prevention and Control (Our World in Data 2020f; 2020e). As of 24 April, there have been 101,790 confirmed cases, of which 2,491 have died (WHO 2020af).

On 24 January, thermal cameras were installed at the Istanbul airports. During February, all flights from China (Daily Sabah 2020; Hürriyet Daily News 2020) and all flights to and from Iran (Wintour 2020), Italy, South Korea and Iraq (Reuters 2020b) were suspended. The list of countries for which the flight ban was imposed was expanded on 13 March to include Germany, France, Spain, Norway, Denmark, Belgium, Sweden, Austria and the Netherlands (Zontur 2020). At the beginning of March, the government recommended that the public as much as possible refrain from going outside during the coming weeks (WKO Austria 2020d). As of 16 March all public facilities, such as schools, universities, theatres, cinemas, bars, and sport centres, have been closed, with the exception of restaurants and shopping centres (WKO Austria 2020d; Habertürk 2020). On 3 April, a 15-day entry ban to 30 Turkish provinces was introduced. Masks covering the mouth and nose became mandatory (CNN Türk 2020). On 11 April, a 48-hour curfew over the weekend was implemented in Turkey’s 31 largest cities, including Ankara, Antalya, Istanbul, and Izmir (Kleine Zeitung 2020b; Hörtenhuber 2020). The weekend curfew was extended for the following weeks (Hörtenhuber 2020).

#### Analysis results

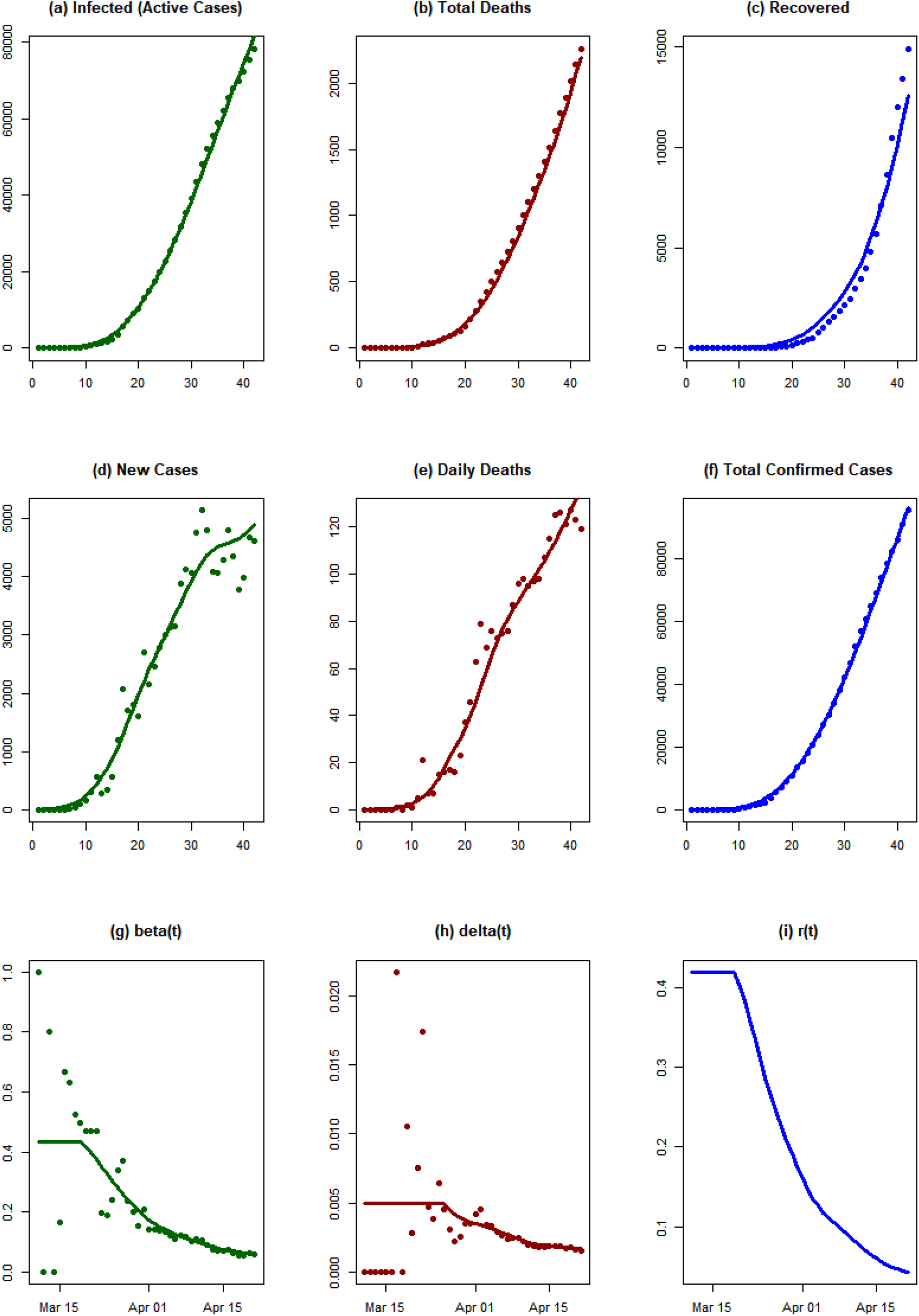

#### Conclusions

Turkey started with a very high infection rate in March 2020. The exponential growth rate of the epidemic has now decreased, as of 21 April. This may be related to various preventive measures taken by the Turkish government, such as travel and gathering restrictions from 13 March, a ban on prayer gatherings in mosques from 16 March, and other social distancing measures. However, the exponential growth rate is still well above the zero line, which indicates that infection is still spreading rapidly.

### Malaysia

#### Background

The first confirmed case in Malaysia was reported on 25 January 2020 (Reuters 2020a). On 4 February 10 cases were confirmed, 7 of whom had a history of travel to China [2]. On 1 March there were 24 infections and no deaths (WHO 2020j), but by 31 March the number of infected people had increased significantly: there were 2,626 confirmed cases and 37 deaths (WHO 2020y). As of 9 April, a total of 4,119 confirmed cases and 65 deaths had been reported to the WHO (WHO 2020ad).

An event held by the Tablighi Jamaat religious movement in Kuala Lumpur is believed to have been an infection hotspot, with about 16,000 people attending from 27 February to 1 March. Two-thirds of the 673 cases confirmed by 17 March were due to this event (Barker 2020).

In February, 3.5 million free face masks were distributed nationwide to control the spread of Covid-19 (The Star 2020).

The travel restrictions imposed by the Malaysian government can be divided into four phases:

1. Early February (09.02.2020): travelers from China (i.e. from Wuhan, Hubei, Zhejiang and Jiangsu provinces) are no longer allowed to enter Malaysia (Borneo Post online 2020);
2. From 13 March onwards, all Malaysians returning from Iran, Italy or South Korea must spend 14 days in self-quarantine in their homes (New Straits Times 2020a);
3. Malaysia bans travelers from Denmark from entering the country as of 14 March (Pei 2020); and
4. On 18 March, all entry to and exit from Malaysia is prohibited (Hassan 2020).

Between 18 and 31 March, controls on movement and social distancing measures were introduced and were then extended until 28 April (Sukumaran 2020; Bunyan 2020; Rodzi 2020). These measures included the cancellation of mass events, closure of kindergartens, state and private schools, and the closure of non-essential businesses (with the exception of food supermarkets and other vital services, such as supplying the population with water and electricity) (Bunyan 2020; New Straits Times 2020b). On 8 April, 24.62 million free face masks were distributed nationwide (Yiswaree 2020).

#### Analysis results

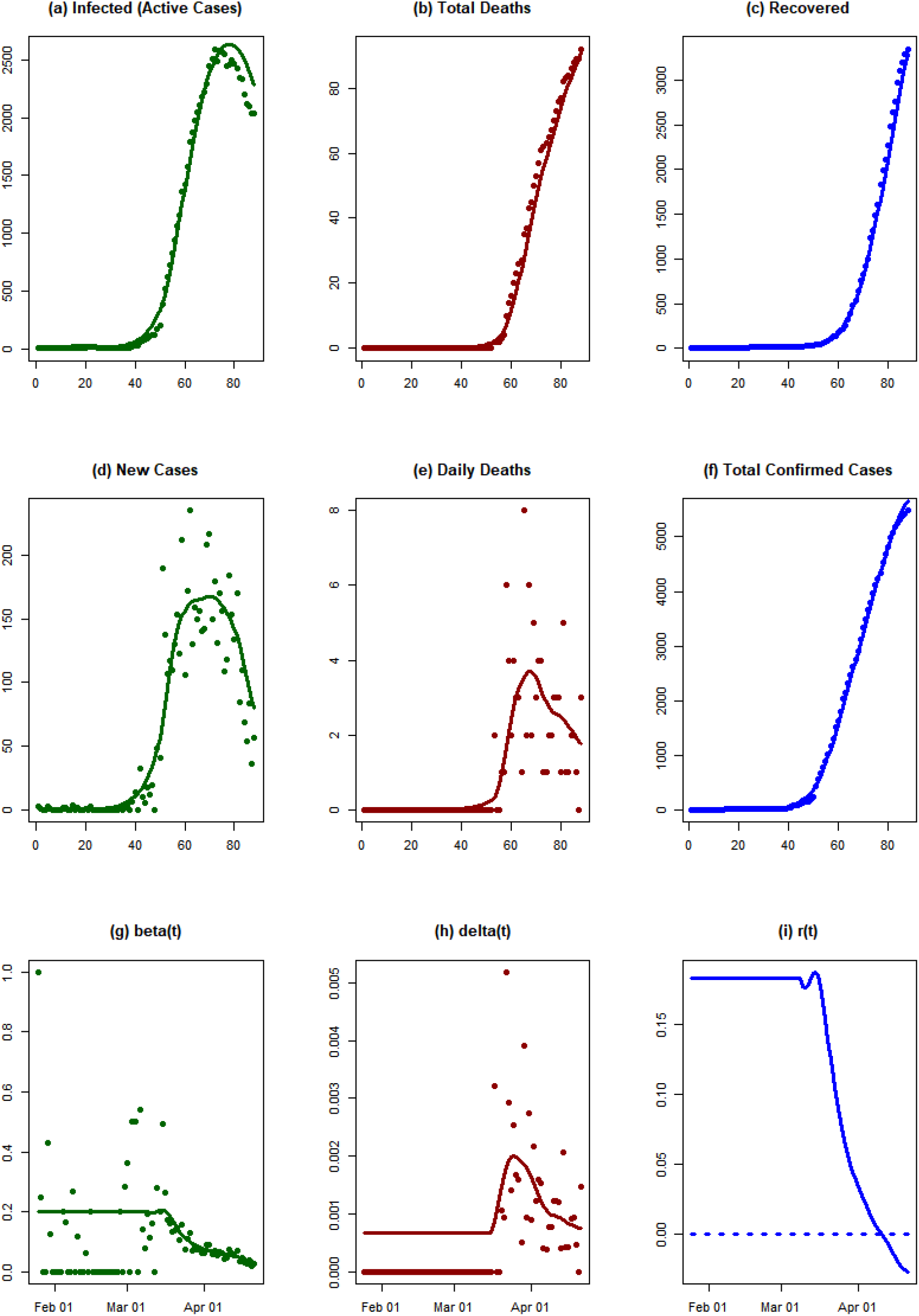

#### Conclusions

The exponential growth rate of the epidemic decreased rapidly in Malaysia from the middle of March 2020. This decay might be related to the movement control order of the Malaysian government, which was implemented on 18 March and extended until 28 April. As of 21 April, the exponential growth rate has crossed the zero line. This shows that the epidemic is dying out.

### Brazil

#### Background

The first confirmed case of Covid-19 infection in Brazil was reported to the WHO on 26 February (WHO 2020o). On 17 March, the Brazilian authorities partially closed the country’s border with Venezuela (Reuters 2020f). On 18 March, six municipalities in the state of Rio de Janeiro – Rio de Janeiro, Guapimirim, Niterói, São Gonçalo, Nova Mesquita and Iguaçu – declared a state of emergency to help contain the novel coronavirus (Globo.com 2020a). On 19 March, the first death was reported and there were 291 confirmed infections (WHO 2020t).

By 20 March, many schools and universities had been closed, partly as a precautionary measure, but also because of proven cases of Covid-19 (France 24 2020; Renteria and Moreno 2020). Many states and municipalities have already declared a state of emergency and implemented measures such as event bans and school closures. On 22 March Brazil closed its borders with all neighbouring countries (WKO Austria 2020e). From 30 March, entry to Brazil by air or land was barred to all foreign nationals for 30 days. However, the movement of goods is not affected by the restrictions and border closures. By 10 April, a total of 15,927 cases and 800 deaths had been reported (WHO 2020ae). Currently, only people with severe symptoms are tested for Covid-19 in Brazil (Globo.com 2020b). At present (as of 11 April 2020), the Brazilian government has not yet decided on any Brazil-wide measures such as school closures or bans on events (WKO Austria 2020e).

#### Analysis results

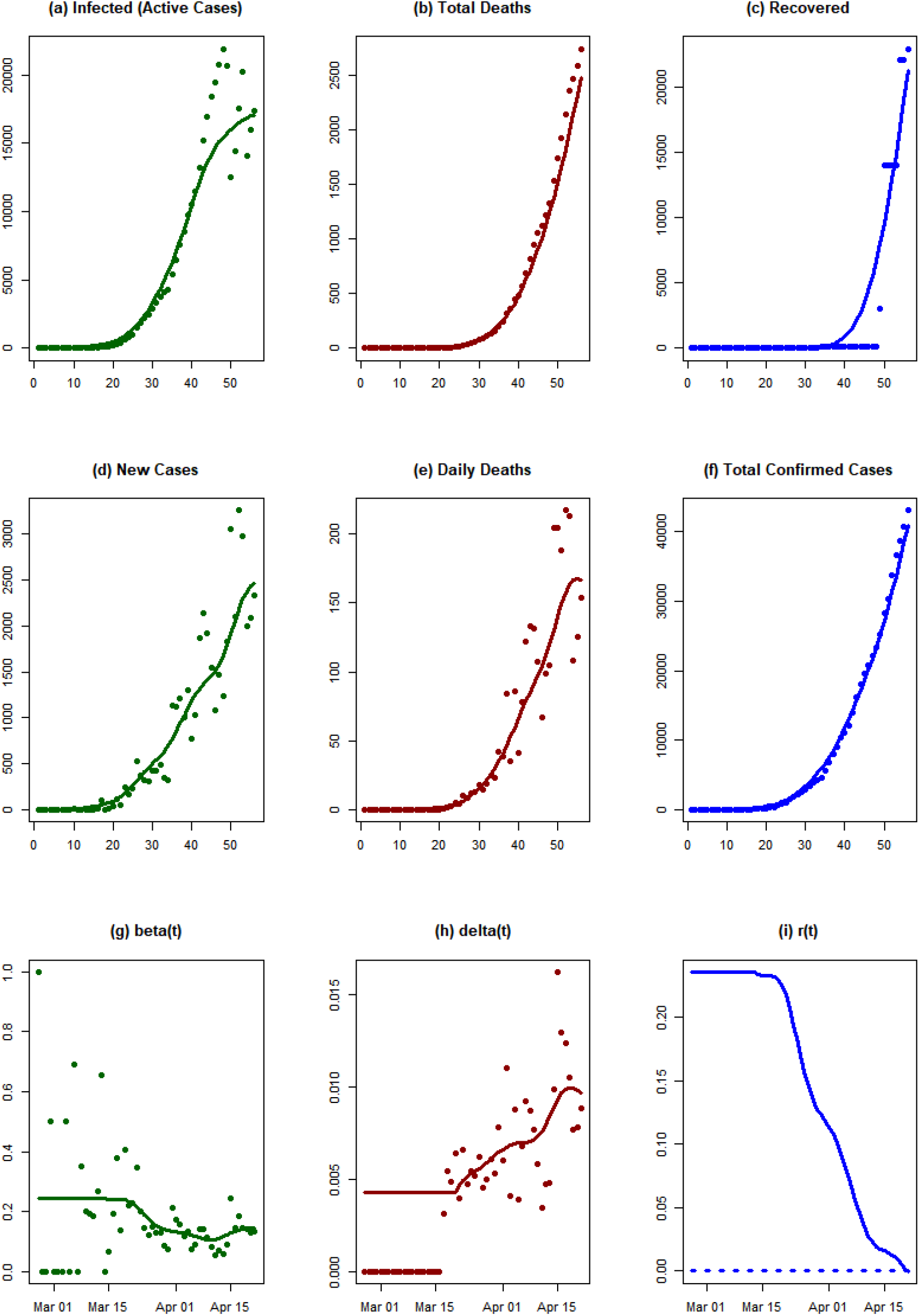

#### Conclusions

We observe a rapid growth in infected individuals during the initial stage of the epidemic in Brazil. As of 21 April 2020, the exponential growth rate of the epidemic has decreased significantly. However, it has not crossed the zero line yet.

### Israel

#### Background

On 21 February, Israel reported its first confirmed case of Covid-19 infection to the WHO (WHO 2020n; Times of Israel 2020a). The first death was reported on 20 March, by which point there was a total of 712 confirmed cases (WHO 2020u). By 10 April there were 9,755 infected and 79 had died (WHO 2020ae). On 26 February, the Israeli government issued a travel warning for Italy and stressed that all trips abroad should be cancelled (YNet News 2020). On 9 March, an obligatory quarantine was imposed on all persons entering Israel: all those entering were ordered to quarantine themselves for 14 days upon entry (BBC News 2020b). The regulation was immediately valid for all Israelis returning home and was applicable to all foreign citizens from 13 March (BBC News 2020b). On 10 March, Israel began to restrict gatherings to 2,000 people. The next day, 11 March, assemblies in Israel were further limited to 100 people. On 14 March, new regulations were announced, which came into force on 15 March. These included a ban on meetings of more than 10 people and the closure of all educational institutions, restaurants, shopping centres, pubs, dance clubs, gymnasiums, venues for events and conferences, cultural institutions, etc. Snack bars, supermarkets and pharmacies were allowed to remain open (Al Jazeera 2020b).

On 15 March, the Israeli government announced that they were considering allowing the Shin Bet internal security service to use mobile phone data to monitor the previous movements of people diagnosed with Covid-19. On 17 March the telephone surveillance program was approved (Gross 2020b; 2020a).

On 1 March, Israel banned all foreigners from entering the country with immediate effect (WKO Austria 2020c). The following day, Prime Minister Netanyahu proclaimed a national state of emergency. On 25 March, more stringent restrictions on the free movement of citizens were imposed by the government. According to the new guidelines, people are not allowed to move further than 100 m from their homes; private vehicles may carry only two passengers; and indispensable workers must be tested for fever at their place of employment. People with a temperature above 38° C are to be sent home (Israel Ministry of Health n.d.; 2020). Since 12 April, all Israelis must cover their nose and mouth when leaving their homes (Times of Israel 2020b).

On 31 March, meetings of over two people (except for close family members) were prohibited (WKO Austria 2020c). On 2 April, the city of Bnei Brak was declared a “restricted zone”, and the Haredi quarters in Jerusalem were locked down on the 12th. According to statistics from the Ministry of Health, almost 75% of coronavirus infections in the city could be linked to these districts (Times of Israel 2020c).

#### Analysis results

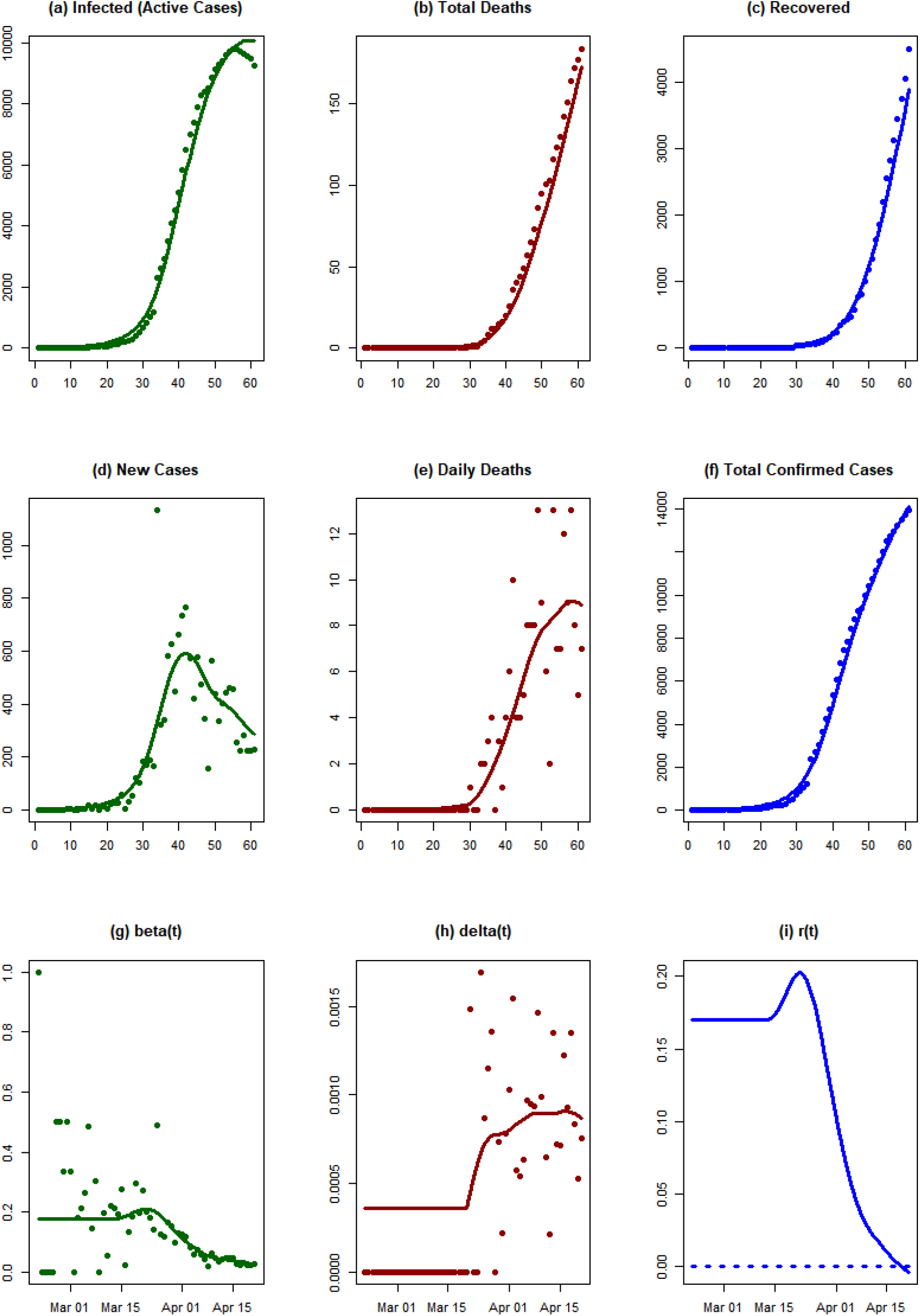

#### Conclusions

We observe a rapid growth in the number of infected individuals between 15 March and 1 April 2020. Subsequently, the exponential growth rate of the epidemic has decreased rapidly. This decay might be related to the strict restrictions on citizens’ movements that were implemented on 25 March. As of 21 April, the exponential growth rate has crossed the zero line, indicating that the epidemic is dying out.

### Philippines

#### Background

The first case of Covid-19 in the Philippines was confirmed on 30 January, in a woman from Wuhan, China (WHO 2020g; ABS-CBN News 2020). By 1 February, two cases and one death had been reported to WHO, but the person who died had been in close contact with the first confirmed case (WHO 2020i). As of 10 April, 4,076 confirmed cases and 203 deaths have been reported in the Philippines (WHO 2020ae).

Since 22 January 2020, tourists from Wuhan have been denied Philippine travel visas under the “Visa-upon-arrival” (VUA) programme (Ramirez 2020). On 2 February, an entry ban was imposed for all foreign travellers who had visited China, Hong Kong or Macao in the previous two weeks (Glee and Gregorio 2020). Holders of permanent visas from the Philippines and Filipino citizens were allowed to enter the country but had to undergo a mandatory 14-day quarantine. On 19 March, the Philippine government announced that it would refuse entry to all foreign nationals “until further notice” (CNN Philippines 2020).

President Duterte issued Proclamation No. 922 on 9 March, declaring a public health emergency (Parrocha 2020). On 13 March it was announced that the National Capital Region (Metropolitan Manila) would be partially locked down for a month (Esguerra 2020); the lockdown was subsequently extended repeatedly and remains in place as of 1 May (Dancel 2020). In addition, local government units outside the National Capital Region were ordered and authorized to impose a local quarantine under certain conditions. On 16 March the province of Luzon was placed under an “enhanced” community quarantine, which includes the following measures: temporary shutdown of public mass transport; restricted land, air and sea transport; a strict house quarantine for all households; and the closure of non-essential businesses (Luna 2020; Office of the President of the Philippines 2020). The Luzon lockdown has been extended until 30 April (PhilStar Global 2020). A general curfew between 8 p.m. and 5 a.m. applies to all residents (WKO Austria 2020b). Other cities and regions have introduced similar measures, including Cebu, Davao City, Ilocos Norte, Ilocos Sur, Iloilo, Negros Occidental, and Negros Oriental (WKO Austria 2020b).

#### Analysis results

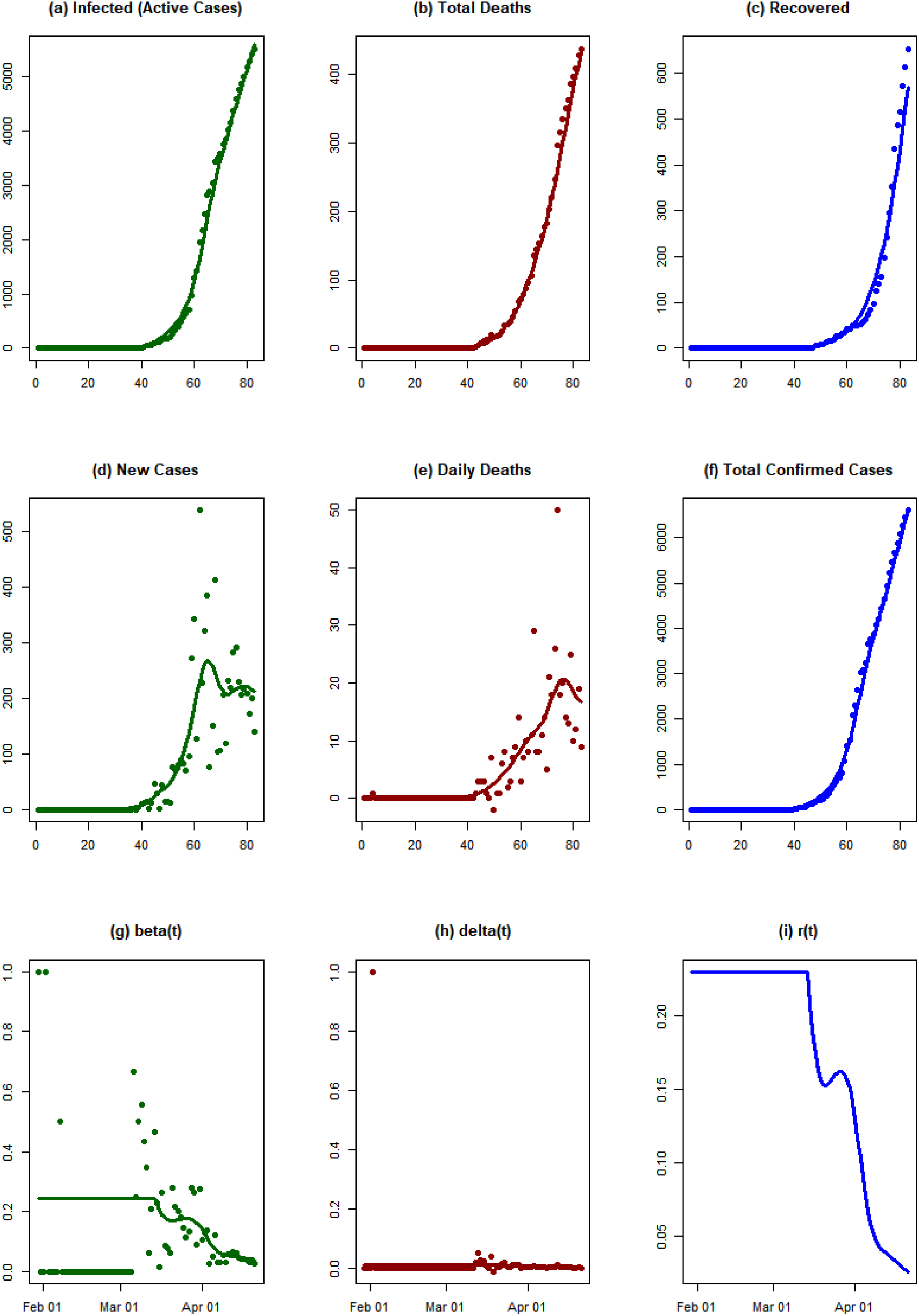

#### Conclusions

The exponential growth rate of epidemic has decreased since the middle of March. This reduction in the rate might be related to various measures taken by the Philippine government, including lockdown measures. However, the exponential growth rate of the epidemic has not crossed the zero line yet.

### Chile

#### Background

On 3 March 2020, Chile notified the WHO of its first case of Covid-19 (WHO 2020r). By one month later (3 April), the number of confirmed cases and deaths had risen to 3,404 and 18 respectively (WHO 2020aa), with the first death reported on 21 March (WHO 2020v). On 10 April the total number of confirmed cases was 5,972 and the number of deaths 57 (WHO 2020ae).

On 13 March, public events involving more than 500 people were banned (Reuters 2020e). One day later, about 1,300 passengers on two cruises in Chilean waters were quarantined because one passenger tested positive for Covid-19 (Roberts 2020). From 15 March to 30 September 2020, cruise ships are no longer allowed to dock in any Chilean port (WKO Austria 2020a). Chile has been tightening its measures and closed its borders for 14 days from 18 March (WKO Austria 2020a). Chileans and foreign nationals who have a permanent residence in Chile can enter the country, but they must agree to a 14-day quarantine upon arrival. Since 16 March schools have been closed. On 18 March the government declared a “state of catastrophe” for 90 days. This was followed by more intervention measures, such as the indefinite closure of all shopping centres, with the exception of essential shops like food supermarkets or pharmacies. On 20 March other businesses, such as restaurants and cinemas, were closed. Two days later, a nationwide daily curfew from 10 p.m. to 5 a.m. was introduced (WKO Austria 2020a).

From 20 March, preventive quarantines were imposed in the regions of Las Condes, La Reina and Vitacura. Since 1 April, health checks have also been carried out at all bus stations, and since 8 April the wearing of face masks has been mandatory on public and private transport (WKO Austria 2020a). Quarantines were extended to certain areas of Chile, e.g. in the cities of Chillán, Osorno, Temuco, Punta Arenas and Padre Las Casas until at least 15 April and to Las Condes until at least 17 April (WKO Austria 2020a).

#### Analysis results

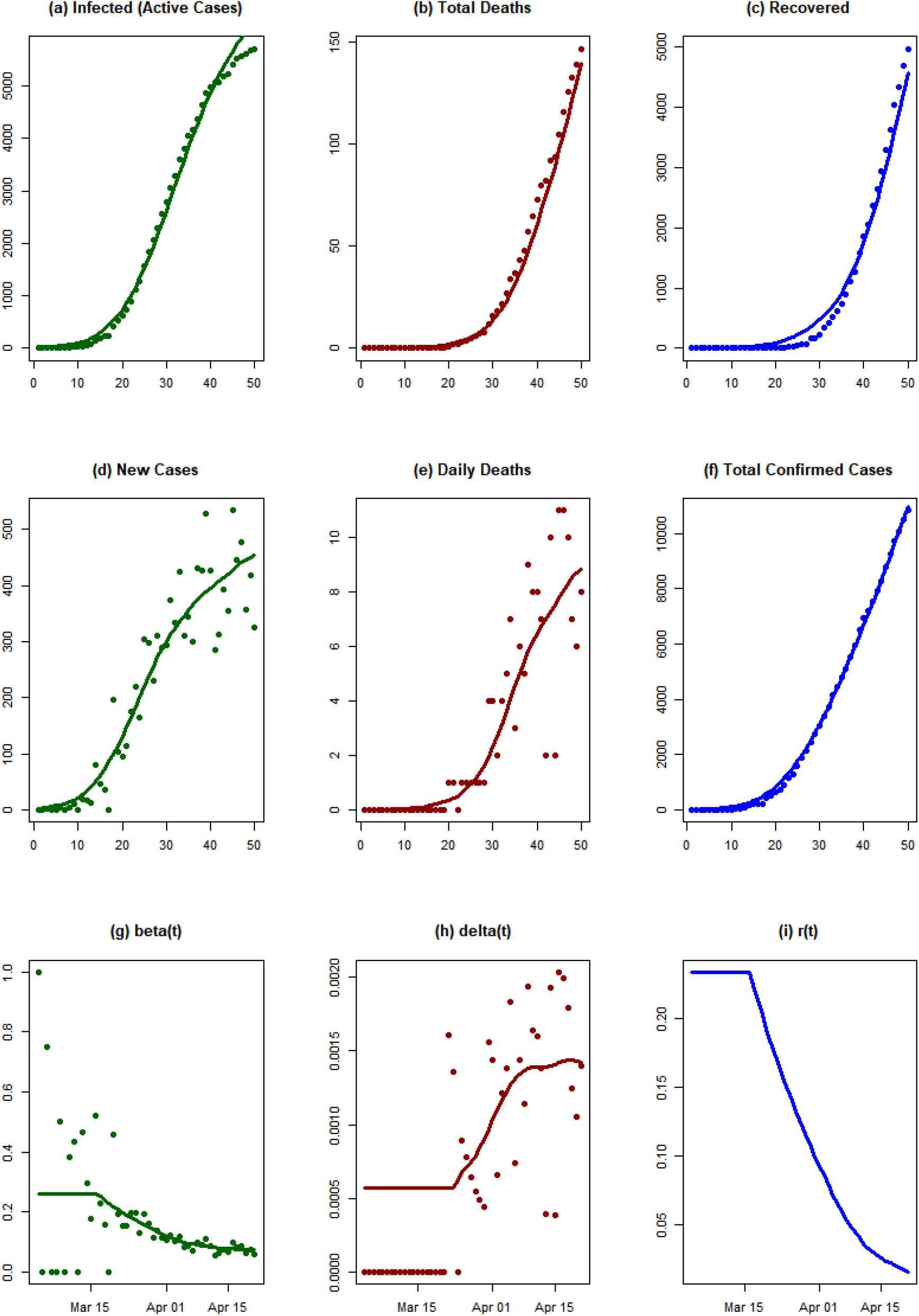

#### Conclusions

The exponential growth rate of the epidemic has decreased gradually from 20 March 2020. This decay might be related to various measures, including lockdown measures taken by the Chilean government during the last week of March. However, the exponential growth rate of the epidemic has not crossed the zero line yet.

### Ecuador

#### Background

The first known case of Covid-19 in Ecuador was in an Ecuadorian woman resident in Spain who imported the virus to the city of Guayaquil on 14 February 2020, showing no symptoms during the flight (El Comercio 2020). This case was confirmed on 29 February, making Ecuador the third country with confirmed cases in South America. The disease spread among the contacts of the first case, leading to 10 confirmed cases by 4 March (CNN Ecuador 2020). On 7 and 10 March, two additional imported cases from the Netherlands were confirmed in the province of Sucumbfos (El Universo 2020c). The first case was also the first confirmed death, on 13 March (Metro Ecuador 2020c). On 14 March the sister of the first case also died of Covid-19 (Metro Ecuador 2020b). The disease spread quickly in the province of Guayas (Expreso Ecuador 2020), where Guayaquil is located. By 5 April, the country had 3,646 confirmed cases and the presence of the disease was confirmed in all 24 provinces (El Universo 2020f). At the time of writing on 13 April, the total count of confirmed cases is 7,466 with 333 deaths (Johns Hopkins University Center for Systems Science and Engineering 2020), making Ecuador one of the countries in South America most affected by the pandemic.

On 26 February, before the first case was confirmed in Ecuador, the government announced measures to prepare for an outbreak, in reaction to the first case reported in Brazil (Secretaría General de Comunicación de la Presidencia 2020a). These measures consisted of airport and seaport checks for travelers coming from affected countries (China, Korea, Italy, and Iran, but not Spain) and the preparation of clinical facilities to handle cases of Covid-19. After the first case was confirmed on 29 February, the government started an active information campaign, including a hotline, online resources, and frequent announcements on social media (Secretaría General de Comunicación de la Presidencia 2020b). Temperature checks and health controls in airports began on 1 March (El Universo 2020a) and the city of Guayaquil applied public social distancing measures and disinfection of public spaces from 7 March (El Universo 2020b). A health emergency was declared on 12 March for a period of 60 days (Metro Ecuador 2020a). From that day, all in-person teaching at schools and universities was cancelled and replaced with online means (El Universo 2020d). Mass gathering events with more than 250 participants were cancelled, and the limit was lowered to 30 participants on 14 March (El Universo 2020e). The same day, air and sea borders were closed and land borders were partially closed, putting travelers into quarantine. On 16 March, a national state of emergency was declared, closing non-essential shops and workplaces, as well as bars and restaurants (El Oriente 2020). On 6 April, additional mobility restrictions were applied and it became compulsory to use gloves and masks in supermarkets (Primicias 2020). The state of health emergency declared can be extended beyond the initially planned period of 60 days and air and sea borders are closed until further notice.

#### Analysis results

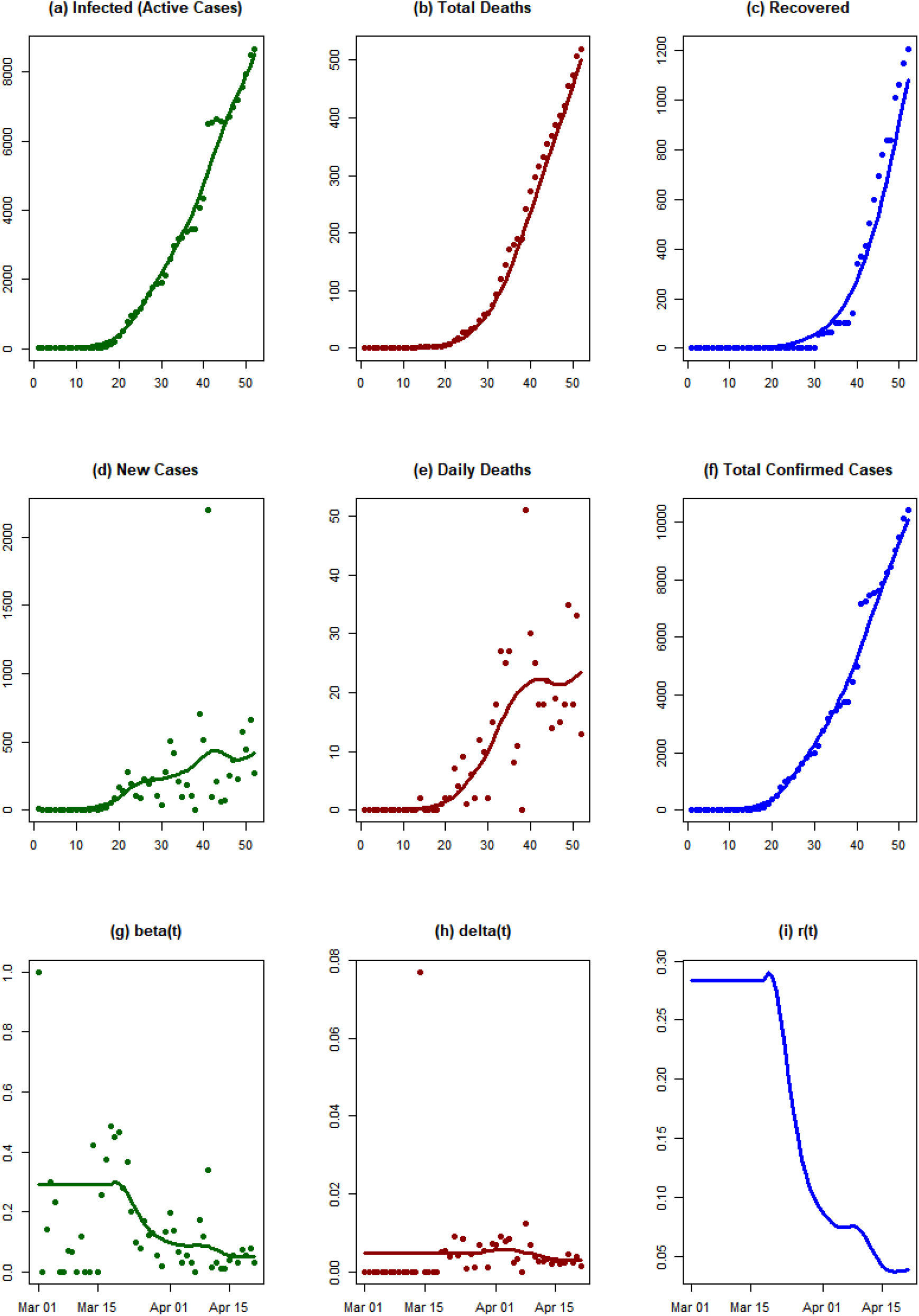

#### Conclusions

The exponential growth rate of the epidemic began to decrease from the third week of March 2020 in Ecuador. This decay might be related to the national state of emergency that was declared on 16 March, closing non-essential shops and workplaces as well as bars and restaurants. However, the exponential growth rate has not crossed the zero line yet, indicating a growth of infected individuals.

### Switzerland

#### Background

The first case of Covid-19 infection in Switzerland was registered on 25 February 2020 in the canton of Tessin: a 70-year-old man returning from Milan (Italy) had contracted the disease (Tagblatt 2020). On 27 February, seven additional cases were reported in the cantons Aargau, Basel, Geneva, Graubünden, Tessin, Waadt and Zurich. All cases were linked to previous stays in Italy. The first death linked to Covid-19 was reported on 4 March, as a 74-year-old woman with a pre-existing condition succumbed to the disease (Der Bundesrat, das Portal der Schweizer Regierung 2020a). At the beginning of March (5 March), Switzerland registered 1.06 confirmed cases per 100,000 inhabitants and was the second most affected country in Europe (CASH 2020). Since 7 March, patients with mild symptoms are no longer tested for Covid-19 (CASH 2020). As of 10 April, 190,000 tests for the virus had been conducted, of which 15% returned a positive result. The median age of infected people in Switzerland is 53 years, and 53% of people infected are women. The death rate is about 2.9% and the median age of people who have died due to Covid-19 is 84 years. Among the deceased, 97% had at least one pre-existing condition (Bundesamt für Gesundheit BAG 2020e).

On 28 February, the Swiss Federal Council declared a *“besondere Lage”* (special situation) (Bundesamt für Gesundheit BAG 2020c), leading to the prohibition of gatherings with more than 1,000 attendees (Bundesamt für Gesundheit BAG 2020b). On 1 March, a large-scale public information campaign was launched, including billboards, flyers and a telephone hotline (Bundesamt für Gesundheit BAG 2020a) and on 11 March nine border crossings to Italy from the canton of Tessin were closed (Eidgenössische Zollverwaltung 2020). On 13 March, the Federal Council implemented several more severe measures, including the prohibition of gatherings of more than 100 people (50 people in restaurants, bars and clubs), and the closure of schools and all skiing areas (Der Bundesrat, das Portal der Schweizer Regierung 2020b). On 16 March, the federal council declared an *“ausserordentliche Lage”* (extraordinary situation) until 19 April, entailing the closure of all bars, restaurants and shops (except grocery stores), the prohibition of all gatherings, and travel restrictions from all neighboring countries except Liechtenstein (Bundesamt für Gesundheit BAG 2020d).

Because of the pronounced federal organization of the Swiss state, several cantons implemented measures at different points in time from the Federal Council. The canton of Tessin has been the most severely affected by the spread of the novel coronavirus because of its proximity to Italy. Therefore it declared a “state of emergency” on 11 March – leading to the closure of all schools and entertainment venues – one week before the Federal Council put similar measures in place (Sansano 2020).

Switzerland has been criticized in the media for its lack of clear and consistent reporting of the number of deaths from Covid-19. Much of the documentation of cases has been carried out on paper rather than digitally, and there have been considerable difficulties keeping track of the actual number of deaths due to the lack of a common reporting standard (Fichter 2020).

#### Analysis results

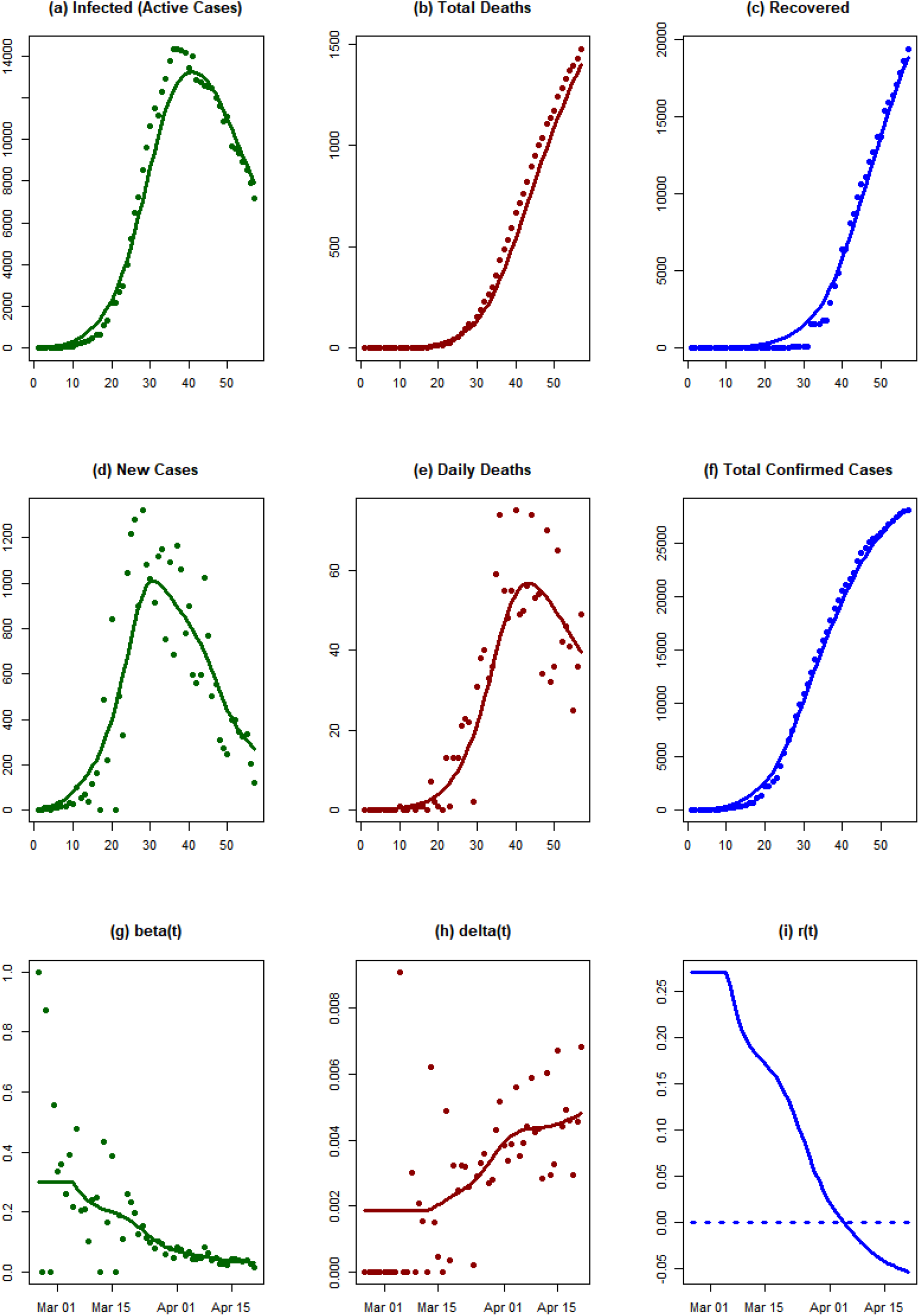

#### Conclusions

Switzerland started with a relatively high exponential growth rate. Subsequently, it has decreased rapidly. As of 21 April 2020, it has crossed the zero line and the epidemic is now dying out.

### Belgium

#### Background

The first case of Covid-19 infection in Belgium was reported on 4 February 2020 (Kockartz 2020), affecting a person returning from Wuhan, China. On 27 February, a second case was reported, in a woman returning from a business trip in northern France (Zeimers 2020). Until mid-March, Belgium registered a rather low number of cases (886 on 15 March), considering that its neighboring countries France and the Netherlands were severely hit by the spread of the novel coronavirus. In late March, case numbers and numbers of deaths started to rise rapidly. As of 10 April, Belgium is reporting 260 deaths per 1 million inhabitants, making it the third most affected country in Europe, behind Italy and Spain (Worldometer 2020a). As of 10 April, there have been 28,018 reported cases and 3,346 deaths, giving a fatality rate of 11.9% (Sciensano 2020). It should be noted that Belgium counts deaths with clinical symptoms of Covid-19 as “suspicious” and includes them in the figures of Covid-19 deaths, even if the patients were not tested (Counasse 2020).

On 10 March, the Belgian government advised that any indoor events with more than 1,000 attendees should be cancelled, but stopped short of a ban (De Standaard 2020a). On 12 March, the government doubled down on the measures and ordered the closure of schools, restaurants, bars, clubs and the cancellation of all public sport and cultural events starting on 13 March (Belgian Government 2020). On 17 March, all non-essential travel was prohibited, all non-essential shops were closed and all gatherings banned (Belgium Official Information and Services 2020). On 20 March the country closed its borders to all non-essential travel (VRT NWS 2020).

Especially in the early phases of the spread of SARS-CoV-2 in Belgium, the Belgian government was criticized heavily by scientists and healthcare organizations for its lack of timely and decisive action to curb the spread of the virus (Wathelet 2020; De Wolf 2020). Belgium’s minister of health failed to secure the supply of medical equipment for the healthcare sector early in the pandemic, as her department’s first order of 5 million masks was not delivered, possibly due to fraud (De Standaard 2020b). In addition, Belgium has been faced with a shortage of reagents needed for testing since the beginning of March, and needed to implement a triage system to limit the number of tests (Hope 2020). The potentially very high “dark number” of people infected with Covid-19 in Belgium due to this lack of testing is a possible explanation for the extremely high fatality rate in this country.

#### Analysis results

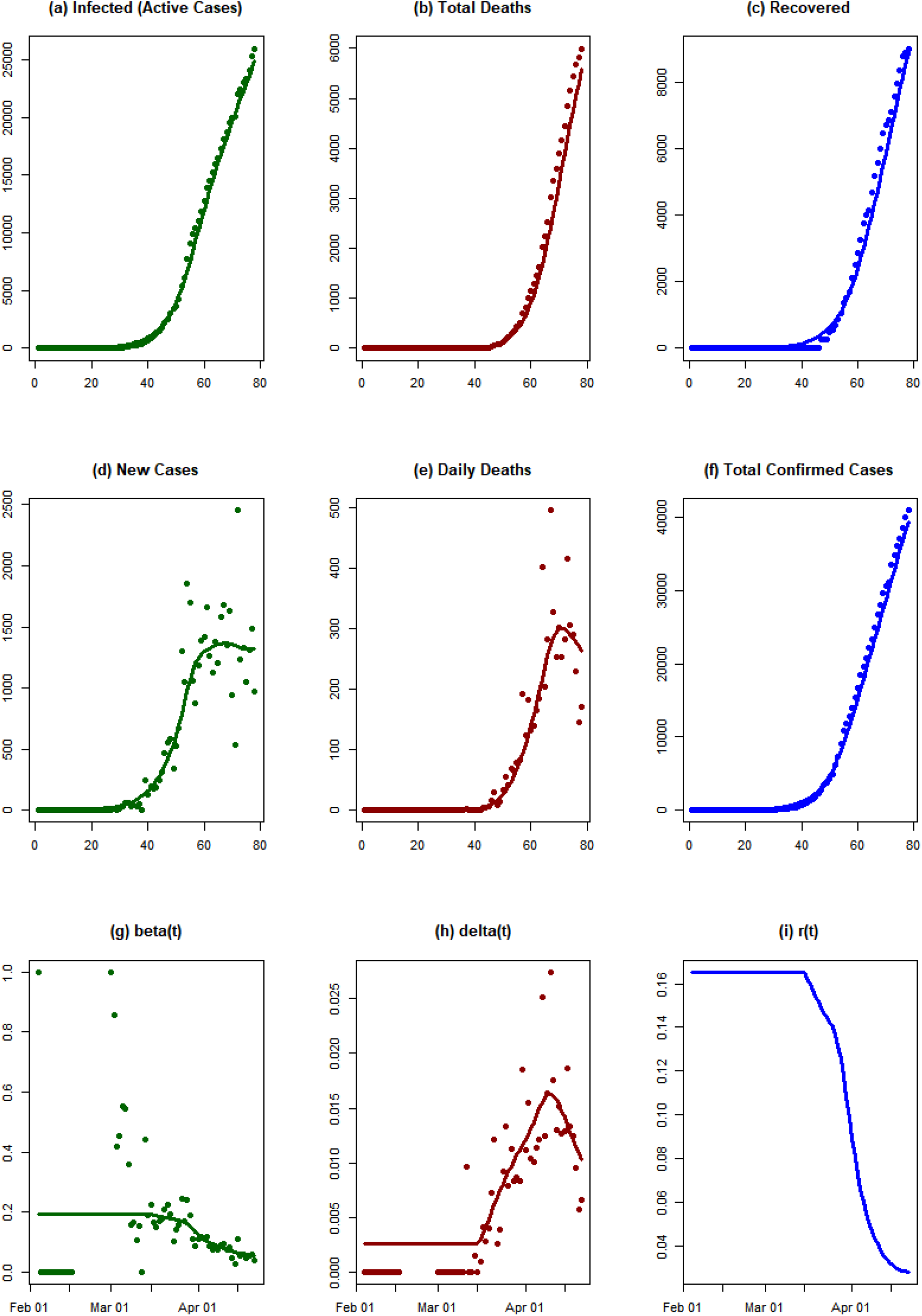

#### Conclusions

The exponential growth rate of the epidemic began to decrease from the middle of March 2020 in Belgium. However, it has not crossed the zero line yet.

### Portugal

#### Background

The first confirmed positive case of Covid-19 infection in Portugal was reported on 2 March 2020, in a man who had returned from a holiday in northern Italy (La Vanguardia 2020). Having reached 78 positive cases (Worldometer 2020b), Portugal declared a nationwide state of alert on 12 March, forbidding large-scale indoor and outdoor events, suspending many international flights, and closing schools and universities as well as bars and restaurants (Franco 2020; OSAC 2020). Nearly a week later, on 18 March, the president declared a state of emergency, to last until at least 17 April (Visit Portugal 2020). The next day, a package of measures was introduced to restrict movement and implement social distancing. Commercial passenger flights were cancelled, public spaces like museums, monuments and tourist attractions were closed, and companies were advised to switch to teleworking (Oliveira 2020). By that time, 642 Covid-19 cases had already been reported (Worldometer 2020b). As the virus was by then spreading within the community and could no longer be contained, Portugal moved into a “mitigation phase” starting from 26 March (Arreigoso 2020). Under the new guidelines, provision of care for those infected with Covid-19 should not be centralized in hospitals, but also include the primary sector (Arreigoso 2020). On 2 April, the Portuguese parliament extended the state of emergency for another 15 days (SAPO 24 2020). Prime Minister António Costa announced the closure of all airports during Easter to slow the spread of the novel coronavirus (Mayberry, Stepansky, and Varshalomidze 2020).

On 5 April, the vast majority out of the total 11,278 confirmed cases in Portugal were recovering at home (9,927), about 9% were hospitalized (1,084) and 267 were in intensive care units (The Portugal News 2020). As of 9 April, Portugal has reached 13,956 confirmed cases, of which 205 have been reported as recovered and 409 have died (Direção-Geral da Saúde 2020a). The majority of confirmed cases (around 8,000) seem to be located in the north of the country, followed by the region of Lisbon with around 3,500. Fewer cases have been observed on the islands of Madeira (53) and the Azores (91). So far, the highest number of new cases on one day (1,035) was recorded on 31 March, with 1,035 (Johns Hopkins University Center for Systems Science and Engineering 2020). Since then, daily increase has lowered and has remained under 852 new cases per day (Direção-Geral da Saúde 2020b).

#### Analysis results

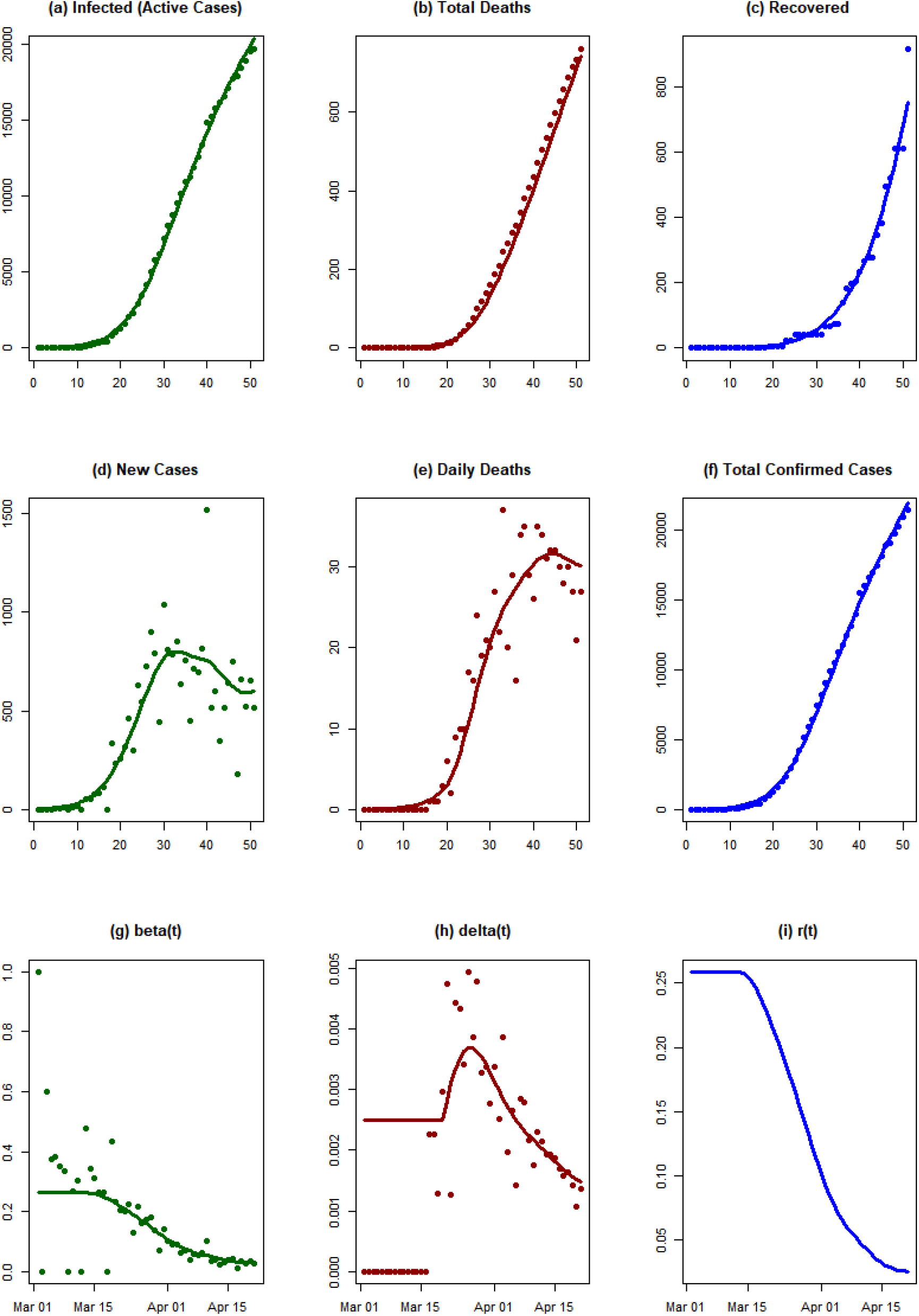

#### Conclusions

Portugal started with a relatively high exponential growth rate. Subsequently, the rate has decreased rapidly, but it has not crossed the zero line yet.

### Russia

#### Background

On 31 January 2020, the first two Covid-19 cases on Russian territory were reported, both in Chinese nationals (TASS Russian News Agency 2020a). A month later, on 2 March, Moscow confirmed its first case, in a man who had returned from a holiday in Italy (Interfax.ru 2020a). The first case of infection not imported from another country was reported on 13 March [3]. The first death of a person who had tested positive for Covid-19 was that of a 79-year-old woman on 19 March. By 31 March, the number of daily new cases had already reached 500, with a majority (387) of them from Moscow (Federal Service for Surveillance on Consumer Rights Protection and Human Wellbeing 2020). As of 10 April, the total number of confirmed cases was 11,917, with 795 recovered and 94 deaths. The number of daily new cases shows no tendency of slowing down (Johns Hopkins University Center for Systems Science and Engineering 2020).

On 23 January, Blagoveshchensk, a Russian city on the border with China, decided to suspend its cultural exchange and official visits to China (Sputnik 2020). Starting on 2 February, the Russian government implemented its first intervention measures. Trains to China were suspended and restrictions on the entry of foreign nationals from China were applied. From 16 February, people returning from China were monitored and ordered to self-isolate, and Chinese citizens were banned from entering Russia. Between 23 and 29 February, Moscow began using a facial recognition network to ensure compliance with quarantine measures. The entry of Iranian citizens was restricted, and the government issued travel warnings for Italy (The Moscow Times 2020b).

Additional measures were implemented in the first week of March. All Russians returning from Covid-19 hotspot countries were ordered to self-isolate; the health ministry recommended against foreign travel; and the export of medical supplies like masks, gloves and protective suits was temporarily banned. Random temperature checks on the Moscow Metro were also introduced (The Moscow Times 2020b). On 11 March, when the WHO declared the virus a pandemic, Moscow banned any events with over 5,000 attendees. Five days later, this restriction was extended to all events with over 50 people. Museums, monuments, circuses and other cultural institutions were closed on 17 March (Meduza 2020). Schools and universities as well as all sports facilities were closed from March 21.

The mayor of Moscow ordered residents older than 65 to self-isolate starting on 26 March, with a reward of 4,000 rubles for following the order (The Moscow Times 2020b). The same day, the ban for foreigners entering Russia was lifted for immediate relatives and spouses of Russian nationals (The Moscow Times 2020b; РБК 2020). Shops, restaurants and bars were closed from 28 March to 5 April in what President Vladimir Putin called a “paid holiday”, and this was subsequently extended to 30 April (The Moscow Times 2020a). On 28 March Russian land and sea borders were closed (The Moscow Times 2020b; Russian Government 2020). From 31 March, Moscow and the surrounding region have enforced self-isolation for all residents, restricting movement to the closest shops and walks within 100 m of their homes (Interfax.ru 2020b). The same restrictions were applied in many other Russian regions during the following days (RIA Novosti 2020; TASS Russian News Agency 2020b; 76.ru 2020). On 9 April, Russia reported that it had carried out more than 1 million tests for Covid-19. Legislation setting out severe punishments, including prison sentences, for those breaking quarantine rules and spreading fake news was signed by President Putin on 10 April (The Moscow Times 2020b).

#### Analysis results

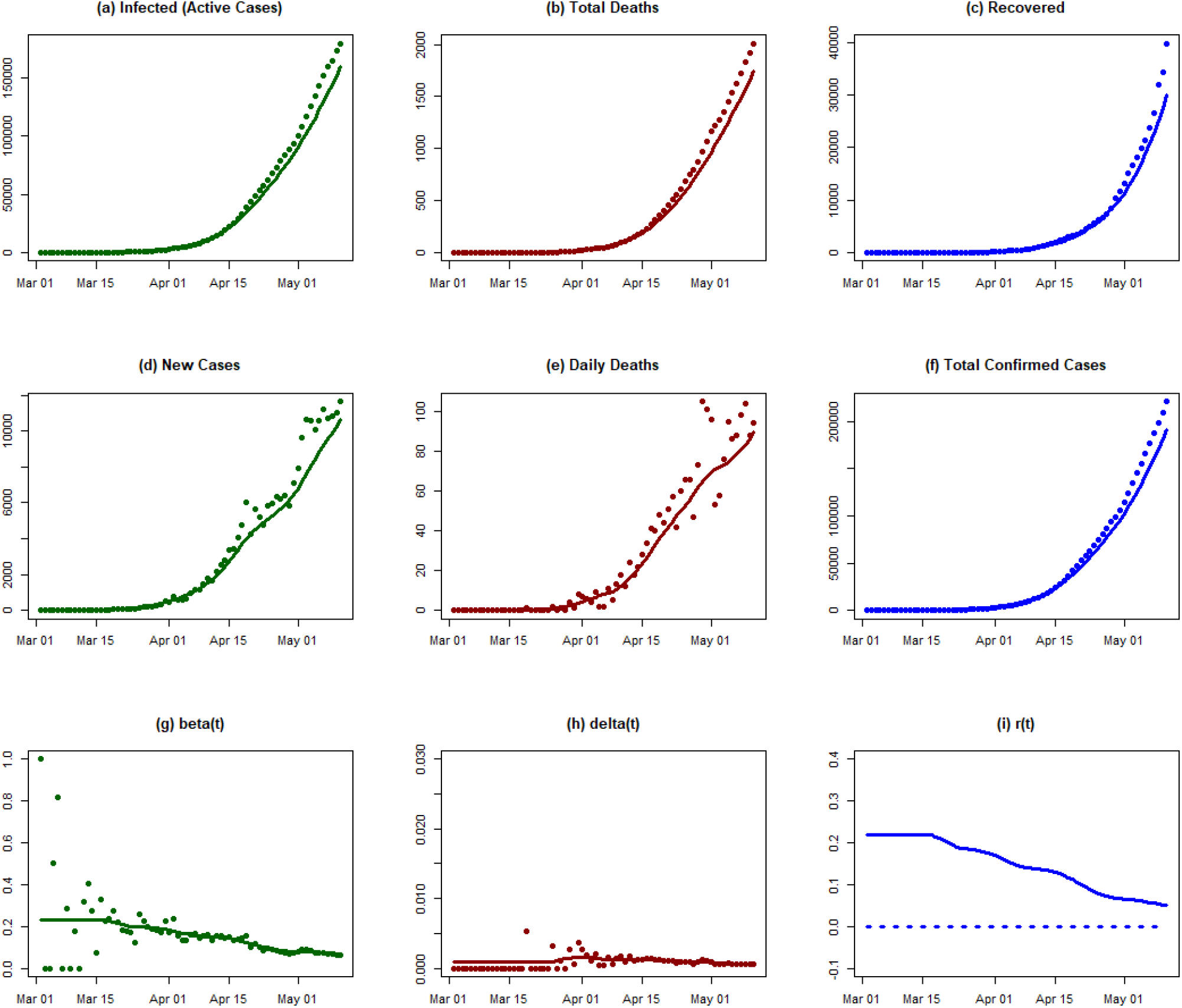

#### Conclusions

We observe a rapid growth in infected individuals from the middle of March 2020 in Russia. The transmission rate has decreased, but very slowly. Between 15 March and 11 May, β has decreased from 0.23 to 0.07, still too high to bring the epidemic under control.

### Norway

#### Background

Norway reported its first case of Covid-19 on 26 February. The patient had returned from China a week earlier (The Local 2020b; Norwegian Institute of Public Health 2020). During the following week, case numbers rose to 56 by 4 March, all of which could either be directly or indirectly (through direct contact with the first patient) linked to foreign travel (Folkehelseinstituttet 2020a). Following the further rise of new cases, the Norwegian government introduced a package of measures on 12 March, largely following the example of other European countries (Nikel 2020). These included the closure of educational institutions; cancellation of sports, cultural, and business events; the closure of bars, restaurants and the entire hospitality sector; and the quarantine of people returning from countries other than Finland or Sweden, regardless of whether they were showing symptoms. Travel within Norway was strongly discouraged (Nikel 2020). On 16 March, further travel restrictions were announced, prohibiting entry to Norway except for residents (Nikel 2020).

The first fatality due to Covid-19 was reported on 12 March, the same day that the first set of measures were introduced (NRK Norge 2020). As of 13 April, a total of 6,488 Covid-19 cases have been confirmed (Folkehelseinstituttet 2020b) with only 134 fatalities, giving a comparably low case-fatality rate (VG 2020).

#### Analysis results

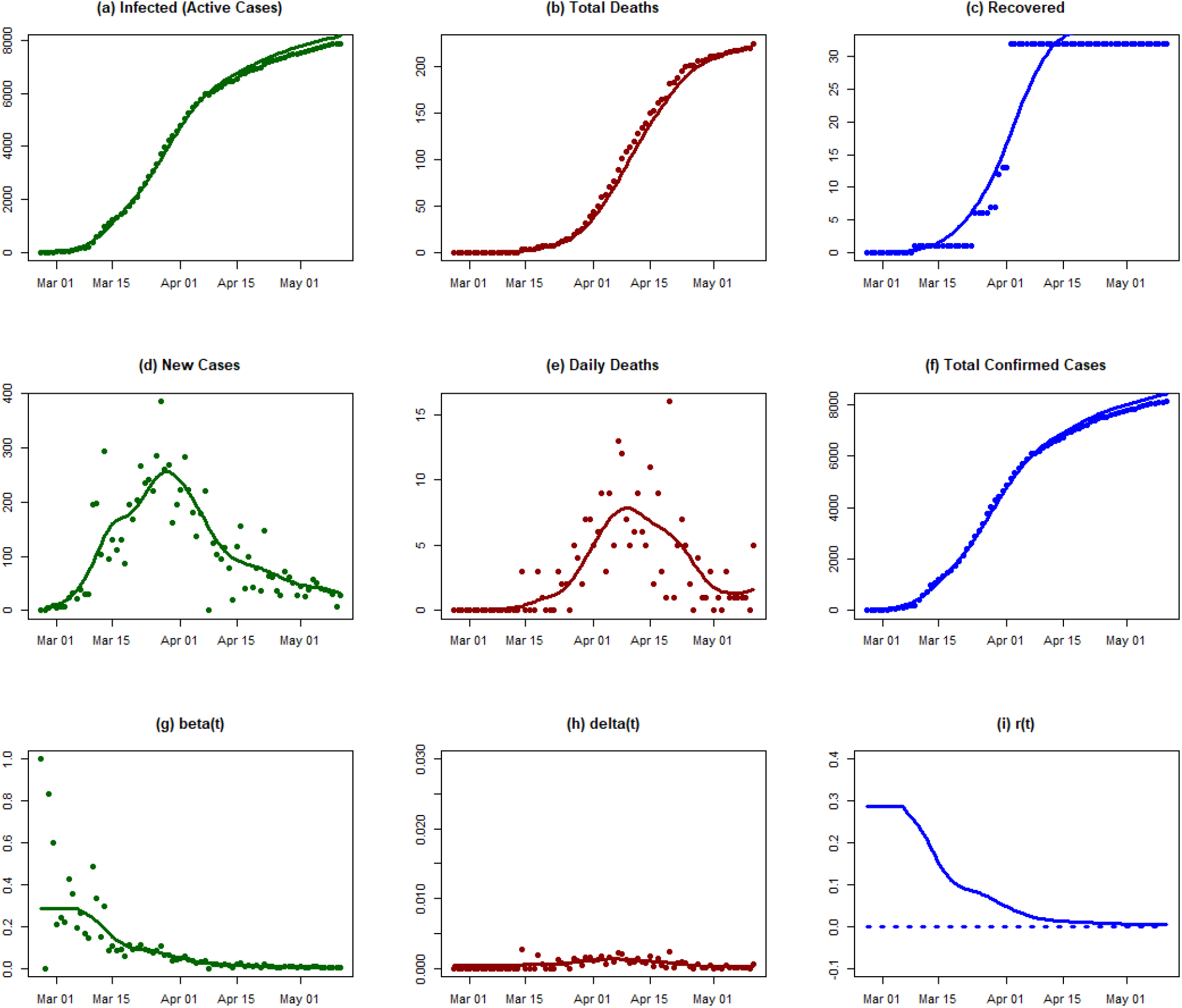

#### Conclusions

The rate of infection in Norway experienced a rapid decline in mid-March followed by more gradual decline thereafter. As of 11 May 2020, the exponential growth rate of the epidemic has not crossed the zero line yet.

### Ireland

#### Background

Ireland reported its first case of Covid-19 on 29 February 2020. The first patient was a student who had recently returned from northern Italy (RTÉ 2020a). Already a month before the first case, Ireland had set up a National Public Health Emergency Team (NPHET), a working group in the Department of Health, to monitor and respond to the outbreak (Boland 2020). A week later, by 7 March, there were 19 cases, the majority of which were independent from one another (Sunderland 2020). Ireland reported its first official death from Covid-19 on 11 March (BBC News 2020d). After that point the number of cases rapidly grew, exceeding 100 on 14 March, 500 on 19 March, and 1,000 on 23 March, according to the Irish government’s online dashboard (Irish Government 2020a). Deaths numbered 2 on 14 March, 3 on 19 March, and 6 on 23 March. As of 6 April there have been 5,364 cases and 174 deaths.

Early attempts to trace the contacts of the first infected individuals led to some school closures (RTÉ 2020a). Already on 6 March, an instance of community transmission of the virus in Cork forced 60 staff at Cork University to self-isolate (RTÉ 2020b). The government introduced measures to control the spread of Covid-19 in stages, similarly to other EU countries. All schools, universities, and childcare facilities were closed on 12 March (BBC News 2020e). St Patrick’s Day events, scheduled for 13-17 of March, were cancelled by 9 March, though some reports suggest pubs were still open and busy until the 14th (BBC News 2020a). Stay-at-home orders and further restrictions on public gatherings were announced on 27 March by Taoiseach (Prime Minister) Leo Varadkar (Irish Government 2020b).

Due to limited testing capacity, Ireland prioritizes the testing of healthcare workers, individuals who have been in direct contact with a positive case, and individuals with a fever and at least one other Covid-19 symptom (Health Service Executive 2020). Some testing facilities are drive-through. Ships of the Irish Navy have been repurposed to test and treat patients (TheJournal.ie 2020). On 6 April the Health Service Executive (HSE) announced that it would double the public health system’s capacity to test within a week, promising a capacity of 4,500 tests per day (RTÉ 2020c).

#### Analysis results

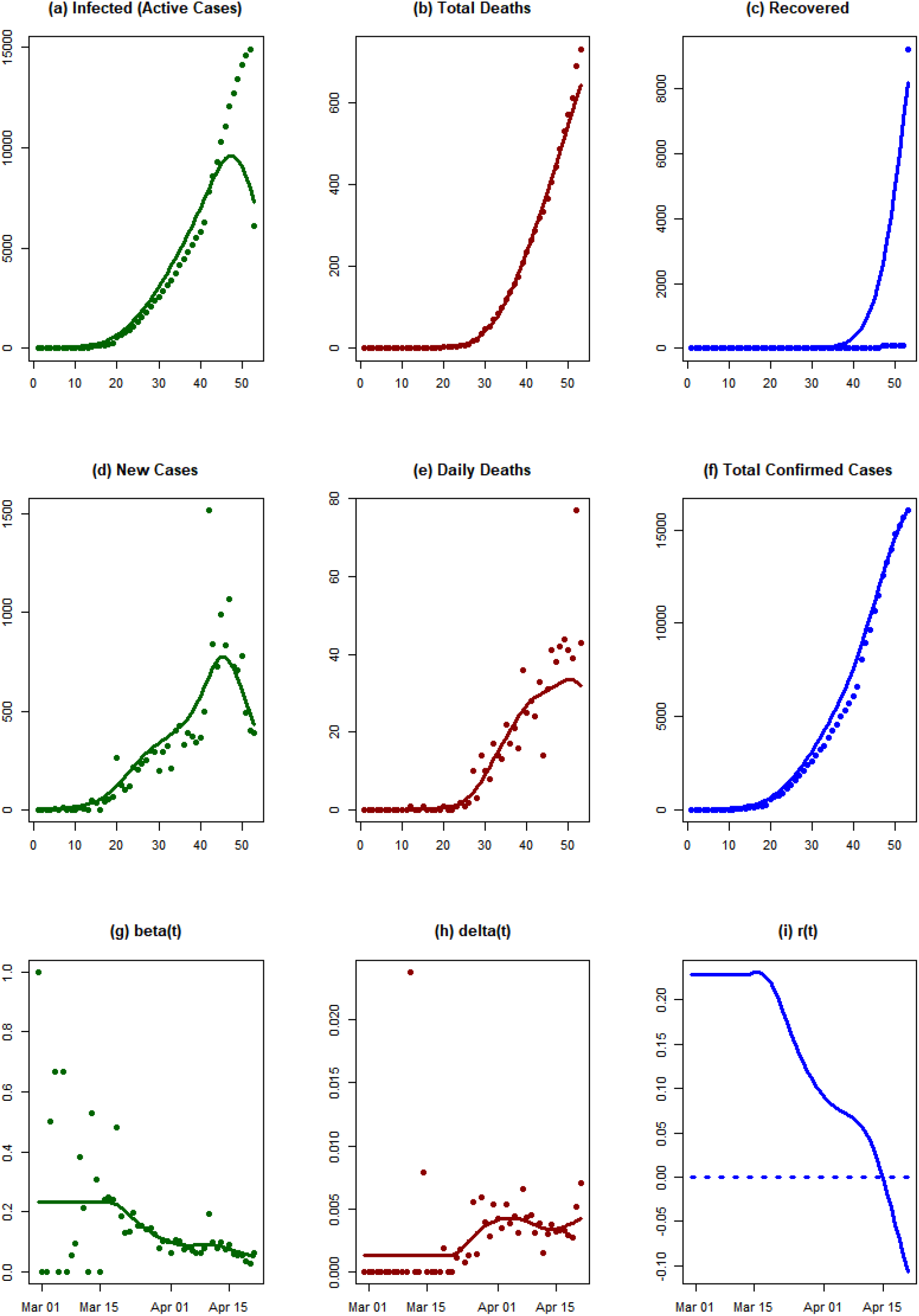

#### Conclusions

As of 21 April 2020, the exponential growth rate of the epidemic is below the zero line in Ireland. The epidemic is dying out in the present environment.

### Mexico

#### Background

The first few confirmed cases of Covid-19 in Mexico were reported on 27 February 2020 (Animal Político 2020a). In a press conference the next day, ministers recommended that the population should practice social distancing and regular hand-washing (Animal Político 2020b). Confirmed cases of Covid-19 grew from 53 on 15 March (Animal Político 2020b) to 118 on 18 March (Animal Político 2020d; 2020c). The first death was reported on 18 March (Animal Político 2020b; 2020c).

Between 20 and 23 March, social distancing measures were imposed, schools were closed and mass gatherings were cancelled (Enciso L. 2020). On 24 March, it was announced that Mexico had entered “Phase 2” of the epidemic, with 5 non-imported cases in the country (Animal Político 2020e). The number of infections grew rapidly, with 475 confirmed cases and 6 deaths as of 25 March (Animal Político 2020f). On 30 March, with 1,093 confirmed cases and 28 deaths, a public health emergency was declared and social-distancing measures were extended until 30 April (Animal Político 2020g).

As of 6 April, 20,475 tests have been carried out; the country has 2,439 confirmed cases, 6,295 suspected cases and has seen 125 deaths linked to Covid-19. The national incidence rate per 100,000 inhabitants is 1.91.

#### Analysis results

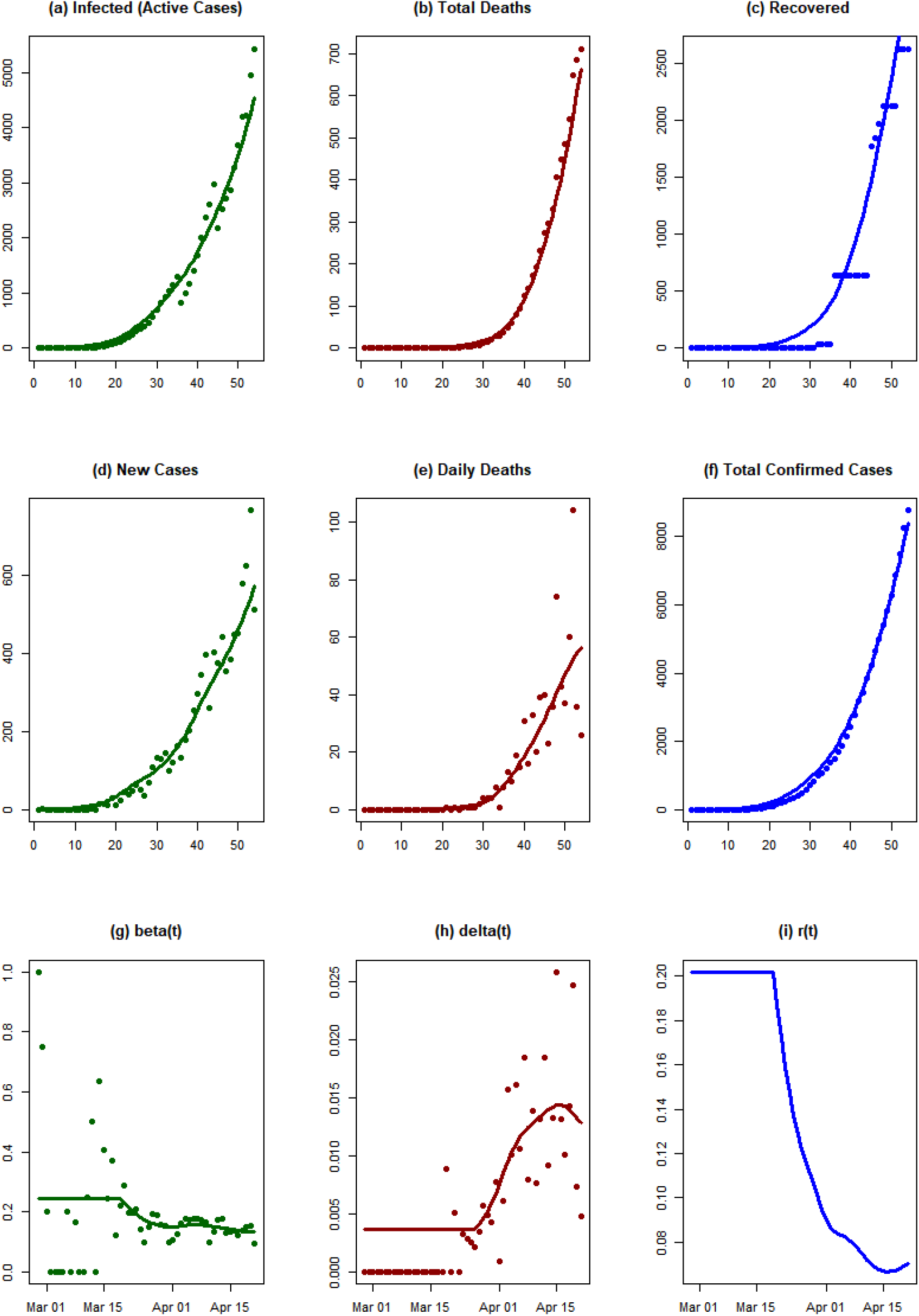

#### Conclusions

We observe a rapid growth in the number of infected individuals from 15 March 2020 in Mexico. As of 21 April, the exponential growth rate of the epidemic has decreased, but it is still well above the zero line.

### Hungary

#### Background

Hungary reported its first two cases of Covid-19 on 4 March 2020 (Reuters 2020c). The first two cases were both Iranian students who had returned to Hungary from Iran in late February to continue their studies. The third case was announced on 5 March, in a British man who had recently visited Milan (Reuters 2020d). By 15 March, 32 cases had been confirmed, many with links to the first two outbreaks (Belügyminisztérium 2020). On the same day, Hungary recorded its first death officially linked to Covid-19. The number of confirmed cases grew quickly, exceeding 100 on 21 March and 500 on 2 April (Átló 2020). As of 9 April, there have been 980 confirmed cases and 66 deaths.

The government established a crisis group with representatives from several ministries in late January (Hír TV 2020). Early steps taken included the return and quarantine of Hungarian citizens living in Wuhan (About Hungary 2020). On 7 March, the government cancelled the national holiday celebrations planned for 15 March (Zoltán 2020). Budapest introduced more frequent cleaning of public transportation and new rules restricting interactions with bus drivers on 11 March (Hungary Today 2020). Also on 11 March, the national government declared a state of emergency, granting itself extra powers to deal with the outbreak (Eder 2020). At this time large gatherings (at least 500 people outdoors or 100 people indoors) were banned, and universities were closed. Travel from Iran, Italy, South Korea and China was restricted. Kindergartens were closed on the 13th, and other schools on the 16th.

On 16 March Prime Minister Viktor Orbán announced further measures, including the closure of national borders to non-citizens (later clarified to exempt some commuters and individuals from the European Economic Area resident in Hungary), the cancellation of all public events, and the daily closure of restaurants by 3 p.m. (Makszimov 2020). A general curfew, restricting movement except for essential activities (grocery shopping, going to the pharmacy, exercise) came into effect on 28 March. Since then, shops have been reserved for those over 65 between the hours of 9 a.m. and 12 p.m.

The Hungarian Parliament has voted to extend the state of emergency indefinitely, allowing the Prime Minister to rule by decree, and banning elections until the parliament lifts the state of emergency. The provisions included the criminalization of the spreading of misinformation, which opposition parties and journalists have claimed could lead to censorship and the suppression of dissent (Makszimov 2020). The decision has drawn international criticism (Rankin 2020).

#### Analysis results

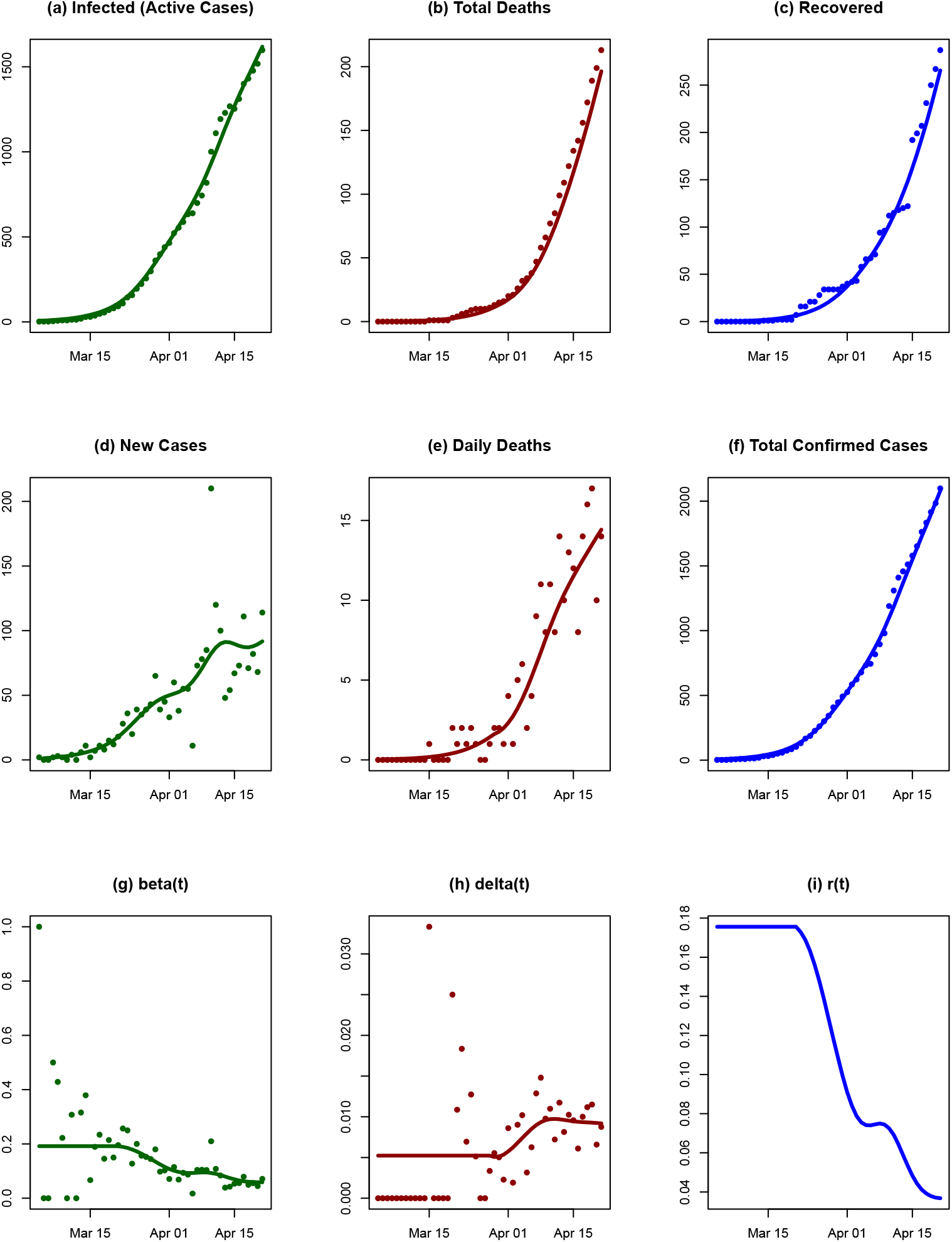

#### Conclusions

The SIRD model fits the data in Hungary well, with mean R2=0.91, particularly the active cases of infection and the total confirmed cases. The fitted model underestimated the total deaths during the observation period. The death rate remains constant at the beginning of until April 1st. The transmission rate decreases gradually around 1 April. The decline in the exponential growth rate of the epidemic may be due to various degrees of social distancing interventions in Hungary, but it has not crossed the zero-line yet, as of 21 April.

### Denmark

#### Background

On 27 February 2020, Denmark notified the WHO of its first case of Covid-19 (WHO 2020o). One month later (27 March), the number of confirmed cases and deaths had risen to 1,877 and 41 respectively (WHO 2020x). On 24 April the total number of confirmed cases was 8,073 and the number of deaths 394 (WHO 2020af).

On 14 March, Denmark closed its borders and public institutions such as schools and universities were closed from 16 March. One day later, gatherings with more than 10 people were prohibited, and these restrictions on the number of people per meeting should continue until 10 May. In the middle of March, restaurants, cafes etc. were also closed. On 15 April, schools opened for specific age groups of children and on 20 April, some businesses, including hairdressers and massage therapists, were allowed to reopen (Pagh-Schlegel 2020; Dagbladet 2020).

#### Analysis results

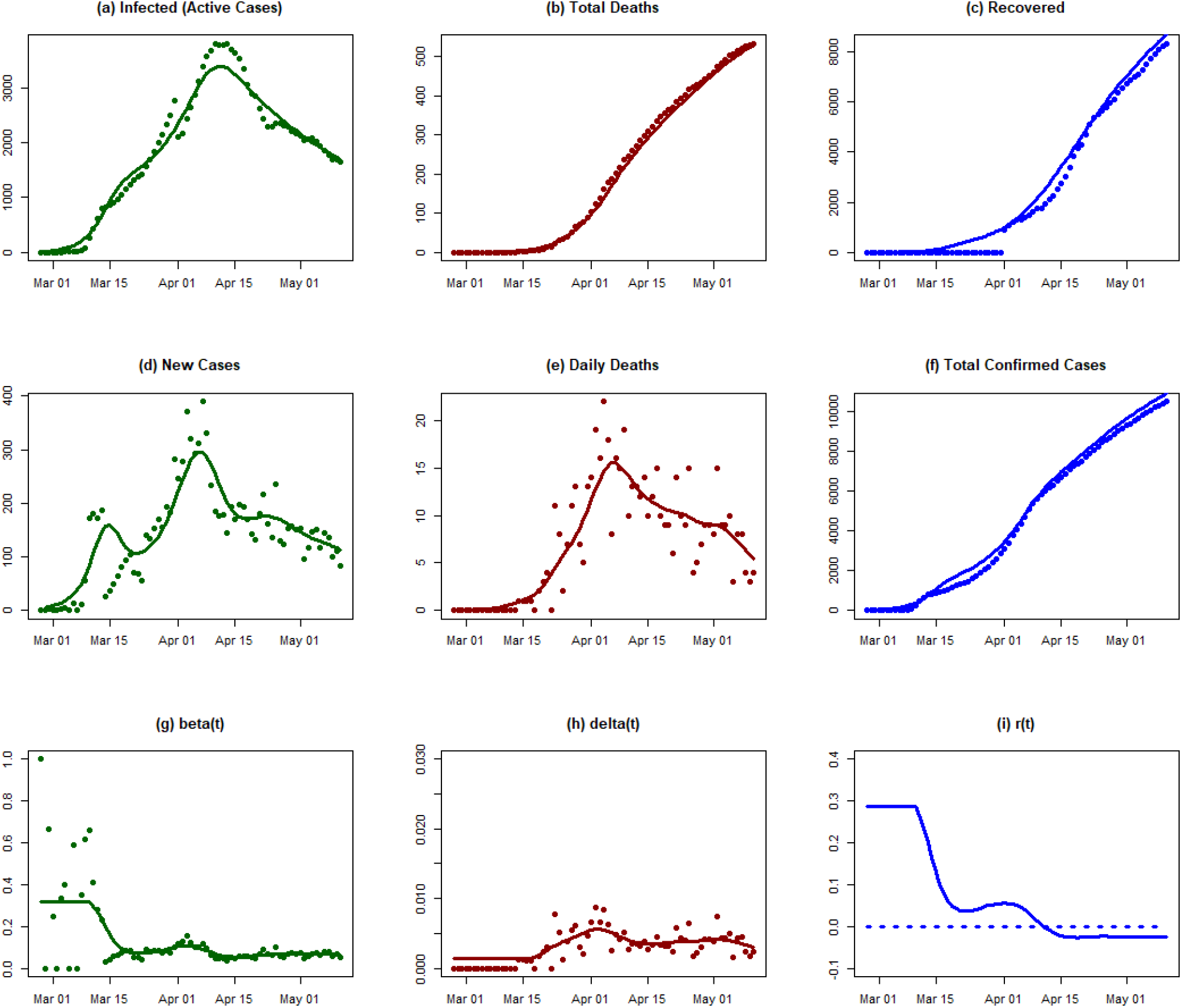

#### Conclusions

The draconian measures implemented by Denmark in mid-March had an immediate effect in bringing an initially high rate of transmission from 0.32 to 0.07. This was, however, followed by a slight rebound. The exponential growth rate crossed the zero line one month later, in mid-April. All indications since then suggest that the epidemic is under control. In particular, partial opening of schools and businesses in late April, has not (as of 11 May) resulted in a second wave of the epidemic.

### Sweden

#### Background

On 31 January 2020, the first case of Covid-19 was confirmed in Sweden, in a woman with a history of travel to Wuhan, China (The Local 2020a). Up to 1 March 2020, 13 confirmed cases had been reported to the WHO (WHO 2020p). As of 24 April, there have been 16,755 cases, of which 2,021 have died (WHO 2020af). On 17 February, the Swedish government recommended against all travel to China’s Hubei province, extending this to Iran on 2 March (Regeringskansliet 2020a; 2020b). Since 19 March non-essential travel to Sweden from countries outside the EU has been prohibited, and on 18 March universities switched to distance learning (WKO Austria 2020f). Further Swedish government recommendations were to practice social distancing and work from home where possible. On 11 March, the government banned all gatherings larger than 500 people, and two weeks later (27 March) reduced this number to 50 people (Krisinformation.se 2020; Die Welt 2020b). Beginning on 1 April, all private visits to nursing homes were prohibited (Lidköpingsnytt 2020). Most restaurants and shops have remained open, though with restrictions designed to enforce physical distancing (Lidköpingsnytt 2020; The Local 2020c).

#### Analysis results

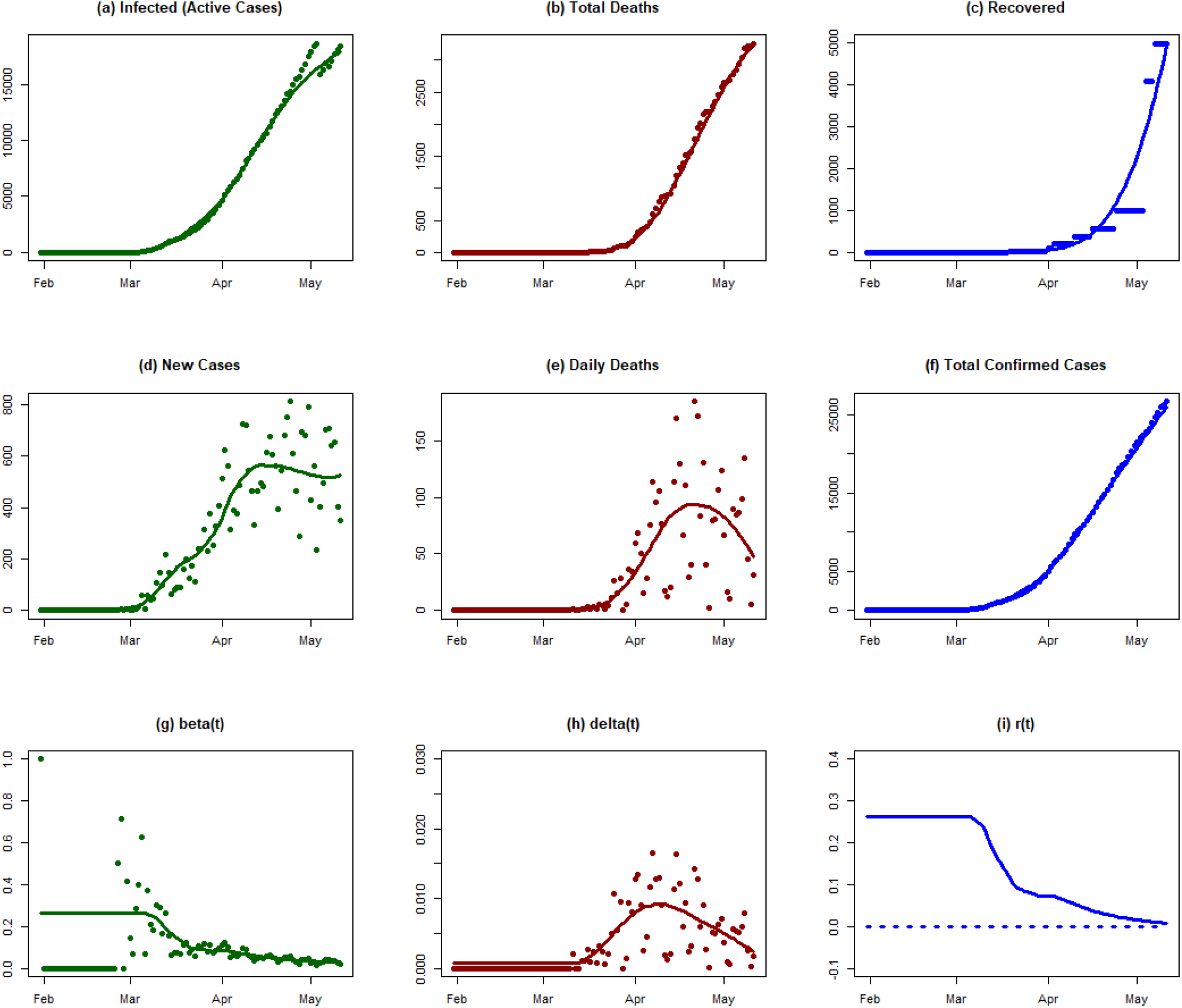

#### Conclusions

Although Sweden did not implement a complete lockdown, as did neighboring Denmark, it was nevertheless able to substantially reduce the rate of transmission in mid-March. Since then, however, the progress has been slow. Unlike Denmark, in which the exponential growth rate crossed the zero line in mid-April, in Sweden r(t) is still positive.

### Germany

#### Background

The first confirmed case in Germany was reported on 28 January 2020 (WHO 2020f). By 1 March, 57 cases had been confirmed (WHO 2020p). On 3 May, a total of 161,703 confirmed cases and 6,575 deaths had been reported to the WHO (WHO 2020ag).

Since 27 February, travelers from China, South Korea, Japan, Iran or Italy have been required to inform the authorities of their whereabouts and of their possible contacts (Der Spiegel 2020a). On 8 March, the government recommended the cancellation of events with more than 1,000 attendees (Tagesschau 2020a). In all federal states, childcare facilities and schools began to be closed between 16 and 18 March (European Union Agency for Fundamental Rights 2020). From 16 March, those wishing to travel from Germany to France, Austria, Luxembourg, Switzerland or Denmark must have special reasons for travelling (Tagesschau 2020b). One day later (17 March) a worldwide travel warning was announced by the Federal Foreign Office (Der Spiegel 2020b).

Several measures based on physical distancing have been implemented in the country, but at different times in the individual federal states of Germany (Dimitriu, Möllers, and Hermann 2020). The first recommendation to avoid social contact was announced on 12 March. On 22 March, the governments of the German states agreed to implement “restriction of social contact” (Bundesregierung 2020a), such as a minimum distance of 1.5 m between people in public spaces. Gatherings were limited to two people, and restaurants were closed (though allowed to serve food and drink to be taken home) (Die Welt 2020a). Mandatory face-mask wearing was introduced at different times in different states: in Saxony on 20 April; in Saxony-Anhalt on 23 April; in Thuringia on 24 April; and in Bavaria, Mecklenburg-West Pomerania and several other states on 27 April (Tagesschau 2020c; Mitteldeutsche Rundfunk 2020; ARD 2020). Some federal states, such as Bavaria, Berlin, Brandenburg, the Saarland, Saxony and Saxony-Anhalt, imposed further restrictions. For example, leaving one’s own home and entering public spaces generally is only possible with a “valid” reason in these states (Mischke 2020; Löbbecke 2020; Ministerin für Soziales, Gesundheit, Integration und Verbraucherschutz 2020).

Some measures were eased from the second half of April. On 20 April, shops with sales areas of up to 800 square metres were allowed to open, though with restrictions on the number of customers allowed in at once. On 4 May, some schools reopened, but mass gatherings are prohibited until at least 31 August. On 30 April, the governments of the federal states agreed to relax further restrictions on public life: museums can open for visitors, church services can take place and playgrounds can be reopened (Bundesregierung 2020b).

#### Analysis results

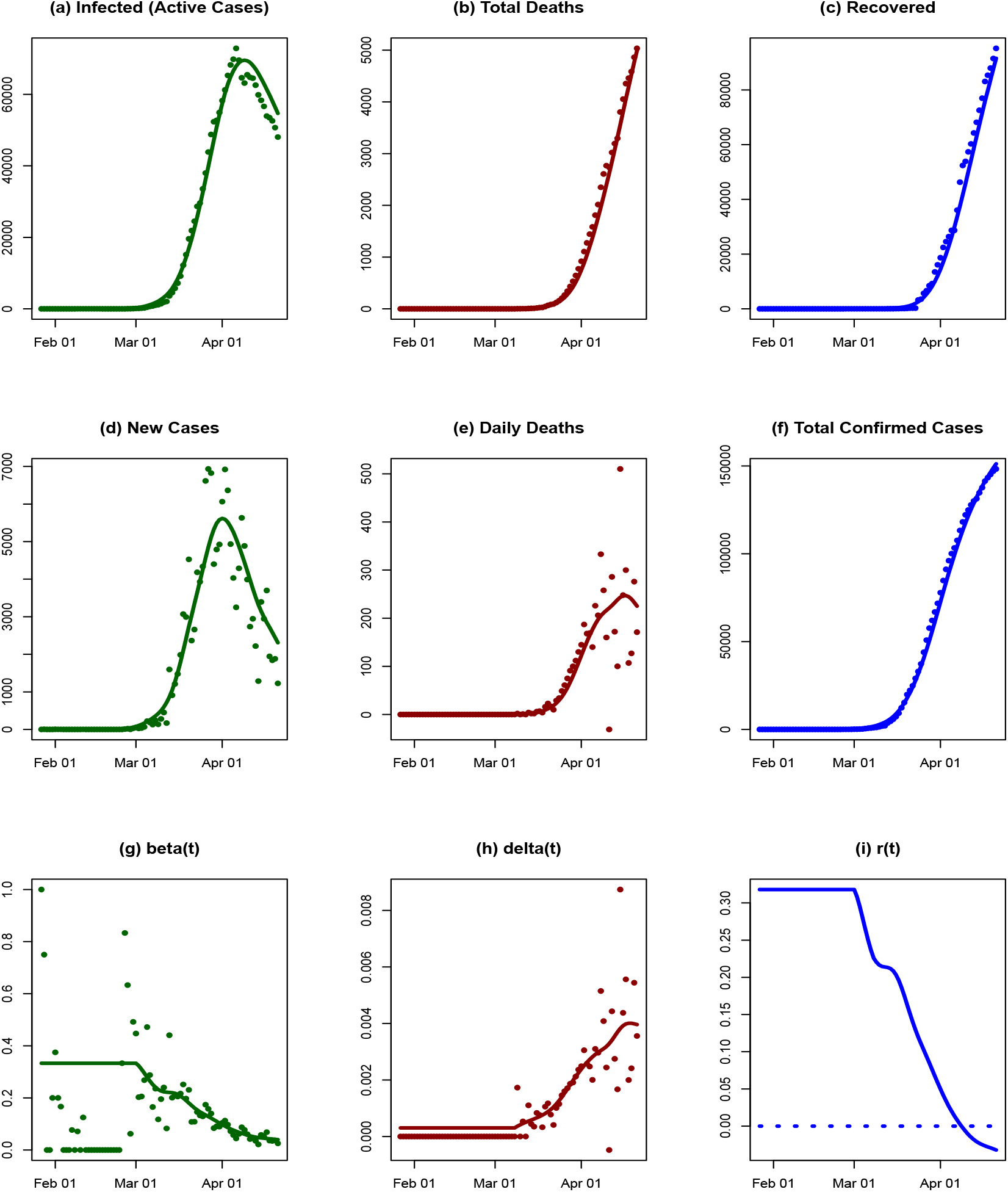

#### Conclusions

The time series consists of a 74-day observation period (27 January-21 April 2020), 14,7065 cumulative confirmed cases, and 4,862 reported deaths. Germany implemented the following measures during the observation period: risk communication, airport health check, resource allocation, increasing hospital capacity, social distancing, and travel restrictions. The number of cases surges around 40 days into the pandemic, which may be due to the increasing testing capacity in the country in early March.

The SIRD model was fitted with a mean R2=0.92. Although the confirmed cases were high in Germany, the death rate was relatively low. The low death rate may be due to the enhanced health capacity, the low mean age of infected patients, and the highly active testing to identify infected patients at an early stage. The model fitted well in terms of total dead and recovered; the number of infected (active cases) was overestimated after the peak (from day 60 onward). The transmission rate declined by about 18% around 1 March, and the exponential growth rate crossed the zero-line within the observation period. The decreasing growth rate suggests that the combination of social distancing and increased testing may have played an important role in mitigating the Covid-19 pandemic in the country.

### Czechia

#### Background

The first three confirmed cases in Czechia were reported on 1 March 2020 (WHO 2020q). One month later (1 April), 3,308 cases and 31 deaths had been confirmed (WHO 2020z). On 3 May, a total of 7,737 confirmed cases and 240 deaths had been reported to the WHO (WHO 2020ag).

On 10 March, schools were closed and two days later the governments declared a state of emergency, mandating the closure of restaurants, bars and non-essential shops (PNP.de 2020; iDNES.cz 2020). On 16 March, Czechia’s borders were closed. Czechia was one of the first European countries to introduce the compulsory wearing of face masks covering the mouth and nose, on 19 March (NTV 2020).

On 14 April, the government presented a plan for returning to normal. For instance, from 20 April, sports facilities and some other businesses were allowed to reopen and weekly farmers’ markets can take place. On 28 April, shops with a floor space under 200 square meters opened. On 11 May, shops up to 1000 square meters may reopen and from 25 May restaurants, bars and other public areas, such as the outdoor areas of zoos, will open (Bućan 2020). On 8 June, all shops will reopen and events with fewer than 50 attendees will be permitted (Němec 2020).

#### Analysis results

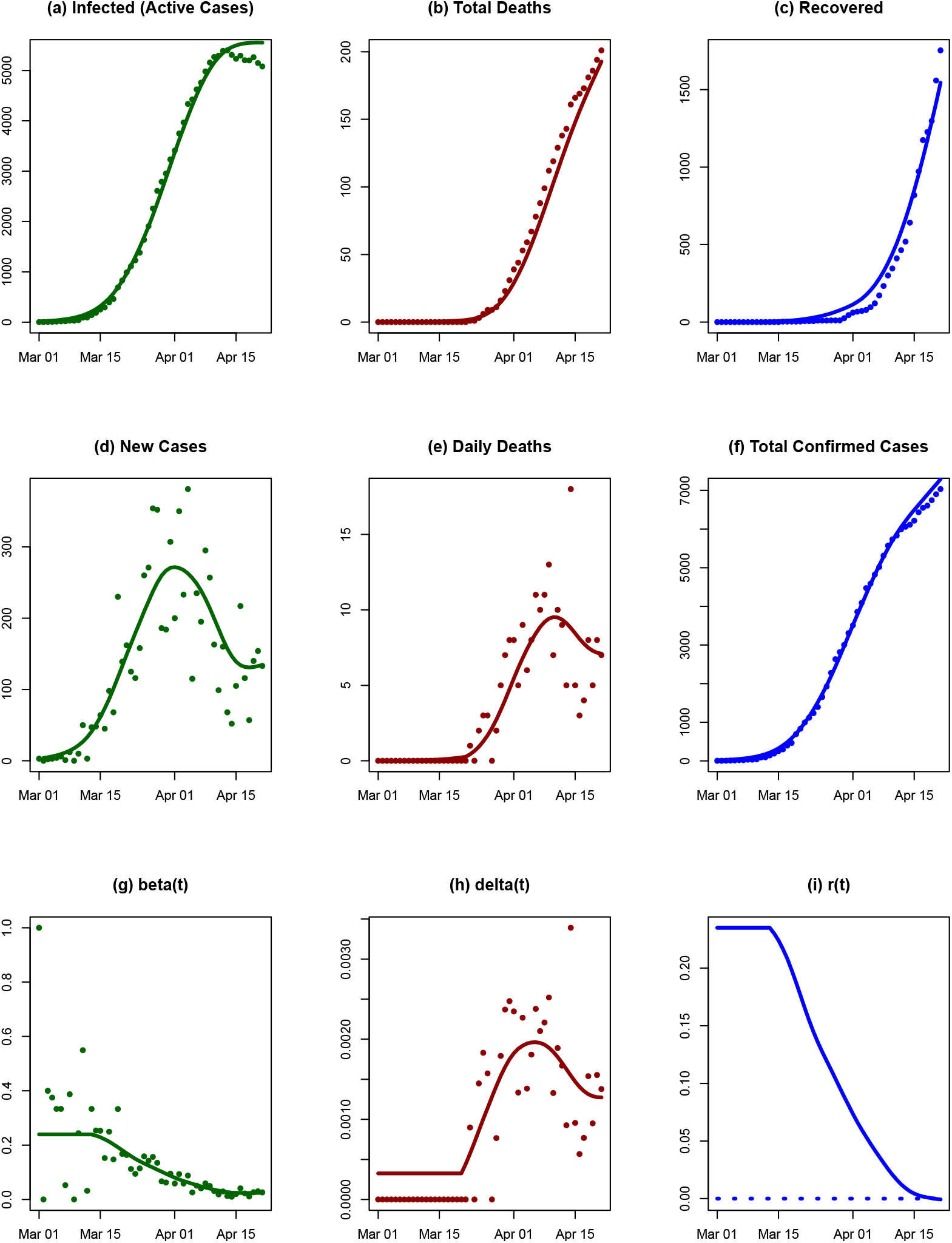

#### Conclusions

The SIRD model fitted the Czech data well, with mean R2=0.90 for an observation period of 52 days. The total dead and recovered were underestimated from day 20 to day 40. The fitted model showed a decrease in new cases from day 30. The transmission rate decreased rapidly from 15 March, by approximately 20%. Czechia implemented lockdown on 16 March (day 16) and mandatory face-mask wearing on 18 March (day 18), which may correspond to the sharpest drop in the transmission rate. The exponential growth rate of disease had almost reached the zero-line by the end of the observation period. The data suggest that the spread of the pandemic is slowing down and is under control in the country.

## Data Availability

We have used the Johns Hopkins University CSSE data repository (Johns Hopkins University Center for Systems Science and Engineering 2020), which provides data on three time-series of Covid-19 indicators: daily counts of confirmed cases (C), recovered cases (R), and deaths (D). 

